# A Genome-wide Association Study Identifies Novel Genetic Variants Associated with Neck or Shoulder Pain in the UK Biobank (N = 441,757)

**DOI:** 10.1101/2024.03.19.24304451

**Authors:** Yiwen Tao, Qi Pan, Tengda Cai, Zen Haut Lu, Mainul Haque, Tania Dottorini, Lesley A Colvin, Blair H Smith, Weihua Meng

**Affiliations:** Nottingham Ningbo China Beacons of Excellence Research and Innovation Institute, University of Nottingham Ningbo China, Ningbo, China, 315100; PAPRSB Institute of Health Sciences, Universiti Brunei Darussalam, Bandar Seri Begawan, BE1410, Brunei Darussalam; School of Mathematical Sciences, University of Nottingham Ningbo China, Ningbo, China, 315100; School of Veterinary Medicine and Science, University of Nottingham, Nottingham, UK, LE12 5RD; Division of Population Health and Genomics, Ninewells Hospital and Medical School, University of Dundee, Dundee, UK, DD2 4BF; Center for Public Health, Faculty of Medicine, Health and Life Sciences, School of Medicine, Dentistry and Biomedical Sciences, Queen’s University Belfast, Belfast, UK BT12 6BA

**Keywords:** Neck or shoulder pain, UK Biobank, genetic correlations, genome-wide association study, genomics, phenome-wide association analysis

## Abstract

Neck and shoulder pain are prevalent musculoskeletal disorders that significantly impact the quality of life for a substantial portion of the global population. Studies have shown that women are more susceptible than men. This study aims to discover genetic variants associated with neck or shoulder pain through a genome-wide association study (GWAS), using data from 441,757 participants in the UK Biobank. The primary GWAS revealed five significant genetic loci (including two novel) associated with neck or shoulder pain, with the most significant single nucleotide polymorphism (SNP) being rs9889282 (*p* = 2.63 x 10^-12^) near *CA10* on chromosome 17. Two novel significant associations were detected on chromosomes 18 and 14, with the top SNPs being rs4608411 (*p* = 8.20 x 10^-9^) near *TCF4* and rs370565192 (*p* = 3.80 x 10^-8^) in *DCAF5*, respectively. The female-specific GWAS identified two significant loci including one near *CA10* and one near *LINC02770* on chromosome 1 with the top SNP being rs5779595 (*p* = 3.57 x 10^-8^). The male-specific GWAS identified one locus in *SLC24A3* on chromosome 20 with the top SNP being rs16980973 (*p* = 6.52 x 10^-9^). The tissue expression analysis revealed a significant association between brain tissues and neck or shoulder pain. In summary, this study has identified novel genetic variants for neck or shoulder pain. Sex stratified GWAS also suggested that gender played a role in the occurrence of the phenotype.

## Introduction

Musculoskeletal pain is a serious concern worldwide, being the second leading cause of disability and with the fourth highest impact on overall health (Vos et al. 2012). Neck and shoulder pain are the second and third most prevalent musculoskeletal diseases, which are major causes of disability (Ferrari and Russell 2003; N. Sanchis et al. 2015). Given that the physical findings and symptoms of neck and shoulder discomfort are similar, these two types of pain are often discussed together (Fish et al. 2011; Meng et al. 2020). The characterization of neck and shoulder pain includes the appearance of stiffness, muscular tension, pressure, or pain in the areas stretching from the neck to the scapular arch (Iizuka et al. 2012).

There is a substantial quantity of epidemiological research indicating that the incidence and prevalence of neck or shoulder pain are high, however this varies widely between studies and nations. The prevalence of neck or shoulder pain in the general health examination program in Norway was 15.4% in men and 24.9% in women, with a sample size of 29,026 (Hasvold and Johnsen 1993). In 2019, the Philippines had one of the highest burdens of neck pain around the world, with an age-standardized point prevalence of 5,333.5 per 100,000 population, and an annual incidence rate of 1,156.3 per 100,000 population (Shin et al. 2022). From a global perspective, the age-standardized median prevalence of neck pain and shoulder pain was 27.0 per 1,000 and 37.8 per 1,000, respectively (Kazeminasab et al. 2022; Lucas et al. 2022). However, because the majority of people do not seek treatment for their problem, it is critical to assess the prevalence of shoulder discomfort in order to examine its influence on the population (Pribicevic 2012). For shoulder problems alone, it has been reported that approximately half of all new episodes that present in medical practice resolve completely within six months, rising to 60% after a year (van der Windt et al. 1996) In addition, frequent recurring neck discomfort not only has a significant influence on an individual’s daily living activities and quality of life, but it also leads to lost productivity and ongoing economic expenditure, resulting in a significant social burden (Kanchanomai et al. 2011).

Several investigations have shown that the occurrence of neck or shoulder pain is significantly related to age, gender, body mass index, physical ability, psychological pressure, workplace and whether suffering from diabetes (Hviid Andersen et al. 2002; Hamberg-van Reenen et al. 2006; Iizuka et al. 2012; Hanvold et al. 2014; Pico-Espinosa et al. 2017). Mahmud et al. (2012) reported that 72% of women and 51% of men suffer from neck pain, indicating that women are more likely to be affected by neck pain than men. Also, the prevalence of neck and shoulder pain in female adolescents increases with age (Hakala et al. 2002). In addition, compared to normal-weight women and men, obese women and men had a 19% and 22% greater risk of neck or shoulder pain, respectively (Nieman 2012). Since neck or shoulder pain is affected by many factors and seriously affects an individual’s physical and mental health, understanding the pathogenesis of neck or shoulder pain is crucial for improving targeted management approaches.

The importance of genetic factors in neck or shoulder pain has been demonstrated. Based on twin studies, environmental and genetic influences on the self-report of neck or shoulder pain have been found (Nyman et al. 2011). It was shown that heritability accounted for 68% of the variance in susceptibility to non-specific neck discomfort at the age of 11-12 years, whereas individual contextual factors accounted for 32% of the diversity in phenotype (Ståhl et al. 2013). Furthermore, the additive genetic effect was twice as strong for concurrent low back pain and neck and shoulder pain as it was for low back pain alone and neck and shoulder pain alone (Nyman et al. 2011). In a previous genome-wide association study (GWAS) using the UK Biobank, the *FOXP2*, *LINC01572*, and *CA10* genes were identified to be significantly associated with neck or shoulder pain, with only *FOXP2* and *LINCO1572* being marginally replicated in another cohort (Meng et al. 2020).

The purpose of the study was to identify novel genetic variants associated with neck or shoulder pain in the UK Biobank by conducting a GWAS on the phenotype using new definitions. Compared with our previous GWAS by Meng et al.(2020), we used a new definition of controls which generated a larger sample size. In addition, we performed new sex-specific GWAS to explore potential genetic variants specifically associated with males or females. We also carried out post GWAS analyses which were not available at the time of the previous publication. Through this study, it was hoped that new genetic associations with neck or shoulder pain could be found, suggesting novel related genetic mechanisms and new targets for the treatment of neck or shoulder pain.

## Methods

### Information about cohorts

The UK BioBank is a unique cohort, facilitating research into health and disease. The cohort is a large-scale collection of broad environmental, lifestyle and genetic data from more than 500,000 volunteers aged 40 to 69 years in the UK from 2006 to 2010. The volunteers gave informed consent to complete detailed questionnaires and provide biological samples including urine, saliva and blood (available at www.ukbiobank.ac.uk). Ethical approval for the study was obtained from the National Health Service and the National Research Ethics Service (reference 11/NW/0382). The genetic data and related data of half a million people in the UK are available for approved research projects into a wide range of diseases.

DNA extraction and quality control (QC) were standardized prior to the release of all data to the UK Biobank, and is described at https://biobank.ctsu.ox.ac.uk/crystal/ukb/docs /genotyping_sample_workflow.pdf. The Wellcome Trust Centre for Human Genetics at Oxford University is responsible for ensuring the reliability of genotyping results. QC steps include identifying poorly performing markers, assessing sample correlations, and accounting for batch effects. These detailed QC steps are available at http://biobank.ctsu.ox.ac.uk/crystal/refer.cgi?id=155580.

### Case and control definitions

A questionnaire was designed by the UK Biobank including a specific pain-related question used in this study: “in the previous month, have you encountered any of the following that interfered with your regular activities?” The alternatives were as follows: (1) Headache; (2) Facial pain; (3) Neck or shoulder pain; (4) Back pain; (5) Neck or shoulder pain; (6) Hip pain; (7) Knee pain; (8) Pain all over the body; (9) None of the above; and (10) Prefer not to say (UK Biobank Questionnaire field ID: 6159). It is possible for volunteers to choose one or more answers. In this study, the cases of neck or shoulder pain were those who answered “yes” to "Neck or shoulder pain". The control group was defined by those who did not choose both " Neck or shoulder pain" and "Prefer not to say". In particular, this study used data from white British descent (field ID: 21000) in order to narrow the population stratification and make the results more specific and interpretable.

### Design of the GWAS

In the primary GWAS, we aimed to understand the genetic basis of neck or shoulder pain. Recognizing the significance of gender in the manifestation of health conditions, our secondary analysis focused on conducting two sex-stratified separate GWAS. This targeted approach in the two sex-specific GWAS aimed to unveil any distinct genetic signatures linked to neck or shoulder pain in males and females.

### GWAS and statistical analysis

Genome-wide complex trait analysis (GCTA, v1.94.1) is a statistical software for estimating the genetic contribution of complex traits across the genome, and is described at https://yanglab.westlake.edu.cn/software/gcta/#Overview (Yang et al. 2011). Utilizing the fastGWA function within GCTA, an ultra-fast mixed linear model association tool was performed for GWAS association analysis by using a sparse genetic relationship matrix. The QC step involved excluding single nucleotide polymorphisms (SNPs) with INFO scores below 0.3, minor allele frequencies lower than 0.5%, and failed Hardy-Weinberg tests (p < 10^-6^). In addition, mitochondrial SNPs and SNPs on the sex chromosomes were excluded. The association tests were adjusted for sex, age, BMI, and eight population principal components using the fastGWAS function in the primary GWAS. Similarly, the association tests were performed with adjustments for age, BMI, and eight population principal components in the secondary GWAS. R v4.2.2 was used to select the data of white British descent in UK Biobank data and divide the cases and controls of neck or shoulder pain. The difference in gender frequency between cases and controls was assessed by a x^2^ test and other covariates were tested by independent t-tests using R v4.2.2, with significance determined by a p-value threshold of 0.05. A threshold of *p* <5 ×10^-8^ is generally considered to be significant across the genome-wide association, which was applied for this study. Furthermore, GCTA was employed to calculate the narrow-sense heritability.

### GWAS-associated analysis by FUMA

The functional mapping and annotation (FUMA) of GWAS web application is an annotation tool for gene analysis, gene-set analysis and tissue expression analysis of GWAS results (Watanabe et al. 2017). The SNP2GENE function accepts as input GWAS summary results and gives thorough functional annotation for all SNPs in genomic regions highlighted by lead SNPs. The number of related loci is ascertained using the "maximum distance of linkage disequilibrium (LD) blocks to merge" method of FUMA, while the number of significant independent SNPs is determined using the r^2^ value (r^2^ > 0.6). In detail, the default FUMA parameters were utilized including the threshold of significant SNPs (*p* <5 ×10^-8^) and the maximum distance of LD (r^2^ > 0.6) with a minimum minor allele frequency (MAF) > 0.01. Also, the maximum distance between LD blocks that might merge into a locus was set to less than 250 kb, and the 1000G Phase3 EUR population was utilized as the reference panel population. Furthermore, Locus Zoom (http://locuszoom.org/) was used to provide regional visualization (Pruim et al. 2010).

The three key aspects of analysis facilitated by FUMA include gene-based association analysis and gene-set analysis conducted by integrating MAGMA (v1.0619) and tissue expression analysis generated by GTEx. In the aspect of gene analysis, the summary statistics of SNPs were integrated at the level of whole genes. Statistical information about the associated SNPs is aggregated at each locus to obtain information about the association with the studied phenotype at the gene level. Integration of the statistical information of SNPs presents the degree of association between each gene and the phenotype under study. All SNPs found inside genes were specifically mapped to 19,023 protein-coding genes and the definition of the significance was p = 0.05/19,023 = 2.60 x 10^-6^. In the gene-set analysis, FUMA performs collective analysis of genomes that share common biological, functional, or other features and the definition of the significance was set at *p* < 0.05/15,485 = 3.23 x 10^-6^. This approach helps to identify specific biological pathways, cellular functions, or other functional groups that further explain the genetic basis of the observed phenotypes. In the tissue expression analysis, FUMA uses tissue-specific expression data from GTEx to analyze the expression level of the specific genes in different tissues.

### Genetic correlation analysis by LDSC

Linkage disequilibrium score regression (LDSC) is a statistical method used to estimate heritability and genetic associations from GWAS summary statistics, which is described at https://github.com/bulik/ldsc (Bulik-Sullivan et al. 2015). LDSC reveals the association between different phenotypes by quantifying the LD between pairs of loci. In addition, LDSC can provide a genome-wide map of genetic correlation, showing the level of genetic correlation of different loci pairs, which helps to identify clusters of loci that are associated with the disease or phenotype as a whole. In this study, LDSC was also used to analyze the genetic correlation of neck or shoulder pain between males and females. The analysis was intended to discover overall genetic difference between male and female patients. We also calculated genetic connections of neck or shoulder discomfort using 1,396 characteristics from the UK Biobank via the Complex Trait Virtual Lab (CTG-VL) (https://genoma.io/). CTG-VL is a complementary and open-source program, which combines available GWAS datasets to facilitate genetic correlation estimation of complex characteristics using LDSC. The results were corrected for multiple testing using the Bonferroni method.

### Phenome-wide association analysis (PheWAS)

We conducted a Phenome-Wide Association Analysis (PheWAS) to explore associations between significant SNP associations and their corresponding genes with other traits with two objectives: (1), aiming to validate GWAS results by confirming the associations we identified with pain phenotypes; and (2) identify novel relationships between significant genetic variants associated with neck or shoulder pain and other phenotypes (focusing on psychiatric phenotypes such as morningness schizophrenia defined by ATLAS). This analysis involved generating PheWAS plots using an extensive dataset of 4,756 GWAS summary statistics available on the GWAS ATLAS platform (Watanabe et al. 2019). For this PheWAS, only SNPs with associations whose *p* values were lower than 0.05 were considered and adjustment for multiple comparisons was carried out using the Bonferroni correction method.

## Results

### Description of the samples

In the initial phase of the UK Biobank study (2006-2010), a cohort of 501,708 participants completed a pain questionnaire. Of these participants, 101,305 reported experiencing ’neck or shoulder pain’, and were thus classified as ’cases’ for the study. In contrast, 340,452 participants, who did not report such pain, were classified as ’controls’. Following the selection of white-British participants and the exclusion of samples failing to meet QC criteria, 98,652 cases (42,869 males and 55,783 females) were included, with 331,541 controls (153,827 males and 177,714 females) in the primary GWAS analysis. This GWAS included 11,165,459 SNPs in the primary analysis. The secondary phase of our study involved sex-stratified GWAS analyses. In this phase, following identical QC procedures, the female cohort (233,497 samples) consisted of 55,784 cases and 177,713 controls, while the male cohort (196,696 samples) included 42,868 cases and 153,828 controls. Table 1 presents a comprehensive overview of the clinical attributes of both the cases and control of the primary and secondary GWAS.

**Table 1.** Clinical characteristics of neck or shoulder pain cases and controls in the UK Biobank.

| GWAS Analysis | Covariates | Cases | Controls | *p*-value |
| --- | --- | --- | --- | --- |
| Primary GWAS | Sex (male: female) | 42,869:55,783 | 153,827:177,714 | <0.001 |
|  | Age (years) | 56.8 (7.95) | 56.9 (8.01) | <0.001 |
|  | BMI (kg/m<sup>3</sup>) | 27.9 (5.00) | 27.3 (4.70) | <0.001 |
| Male-specific GWAS | Age (years) | 57.1 (8.06) | 57.0 (8.09) | <0.05 |
|  | BMI (kg/m<sup>3</sup>) | 28.3 (4.42) | 27.7 (4.19) | <0.001 |
| Female-specific GWAS | Age (years) | 56.7 (7.85) | 56.6 (7.94) | <0.05 |
|  | BMI (kg/m<sup>3</sup>) | 27.6 (5.39) | 26.9 (5.07) | <0.001 |

Total number of females: 233,497; Total number of males: 196,696.
A $\chi^2$ test was used to test the difference of gender frequency between cases and controls and an independent t test was used for other covariates.
Continuous covariates were presented as mean (standard deviation)
BMI represents body mass index.

### GWAS Results

In the primary GWAS, five GWAS signals were identified that demonstrated GWAS significant associations with neck or shoulder pain. These signals reached genome-wide significance with *p* values less than 5 x 10^-8^, as shown in Figure 1. Of these five loci, two are newly identified. Table 2 offers a comprehensive overview of the top SNPs within each loci. For a detailed list of all SNPs that displayed significant associations in this GWAS, please refer to Supplementary Table 1.

**Fig. 1.**
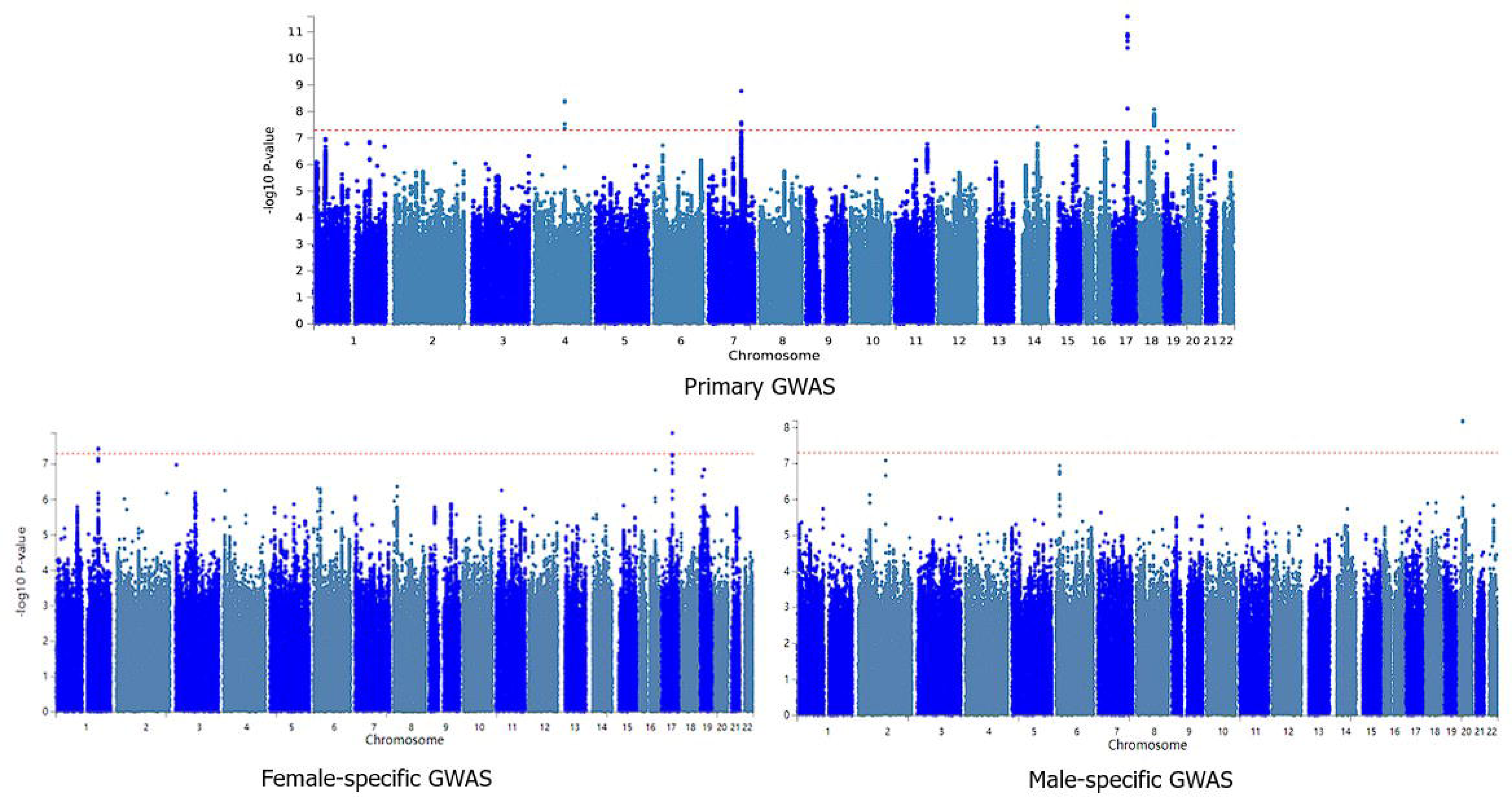
The Manhattan plot of the primary GWAS analysis on neck or shoulder pain (N = 441,757), the female-specific GWAS analysis (N = 239,772) and the male-specific GWAS analysis (N= 201,985) The dashed red line indicates the cut-off *p* value of 5 × 10^−8^

**Table 2.** The top SNPs within 5 loci identified by the GWAS on neck or shoulder pain.

| Locus Rank | rsID | Chr | SNP position | Nearest Gene | Effect allele | Non effective allele | Frequency the effect allele | p value | Beta | Identified or novel |
| --- | --- | --- | --- | --- | --- | --- | --- | --- | --- | --- |
| 1 | rs9889282 | 17 | 50259142 | *CA10* | A | C | 0.61 | $2.63 \times 10^{-12}$ | -0.007 | Meng et al. |
| 2 | rs34291892 | 7 | 114058731 | *FOXP2* | CA | C | 0.62 | $1.69 \times 10^{-9}$ | 0.005 | Meng et al. |
| 3 | rs13135092 | 4 | 103198082 | *SLC39A8* | A | G | 0.92 | $3.84 \times 10^{-9}$ | -0.01 | Tsepilov et al. |
| 4 | rs4608411 | 18 | 52824637 | *TCF4* | C | T | 0.67 | $8.20 \times 10^{-9}$ | -0.005 | novel |
| 5 | rs370565192 | 14 | 69515858 | *DCAF5* | TC | T | 0.63 | $3.80 \times 10^{-8}$ | -0.005 | novel |

Chr: chromosome

The primary GWAS revealed the strongest association in the SNP cluster near the *CA10* gene on chromosome 17q21.33, a *p* value of 2.63 x 10^-12^ for rs9889282. A significant association was also identified in the *FOXP2* gene on chromosome 7, with a lowest *p* value of 1.69 x 10^-9^ for rs34291892. Another notable association was identified in the *SLC39A8* gene on chromosome 4, with a lowest *p* value of 3.84 x 10^-9^ for rs13135092. Additionally, significant associations were detected on chromosomes 18 and 14, with the top SNPs being rs4608411 (*p* = 8.20 x 10^-9^) near *TCF4* and rs370565192 (*p* = 3.80 x 10^-8^) in *DCAF5*, respectively. These loci on chromosomes 18 and 14 are reported here for the first time in association with neck or shoulder pain. The regional plots displaying the most significant locus along with two novel loci are featured in Figures 2 and 3, while the regional plots for the additional two loci are available in Supplementary Figures 1. The Q–Q plot of the GWAS during the discovery phase is depicted in Supplementary Figure 2. The SNP-based heritability for neck or shoulder pain was determined to be 0.05, with a standard error of 0.02.

**Fig. 2.**
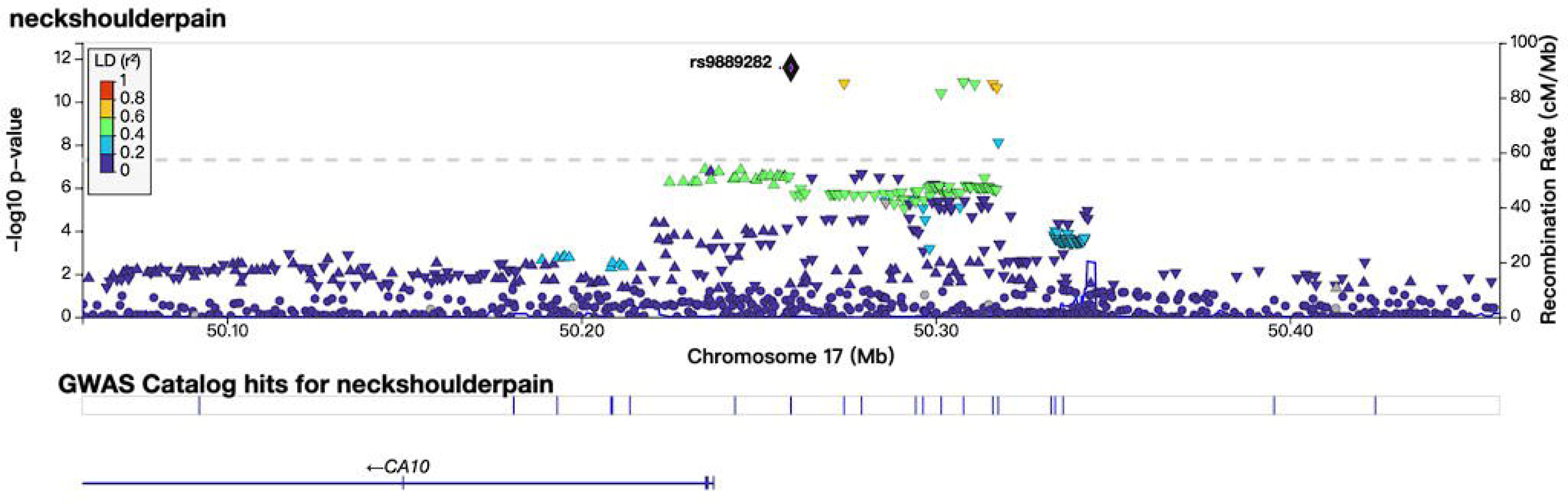
The regional plots of locus in the CA10 region

**Fig. 3.**
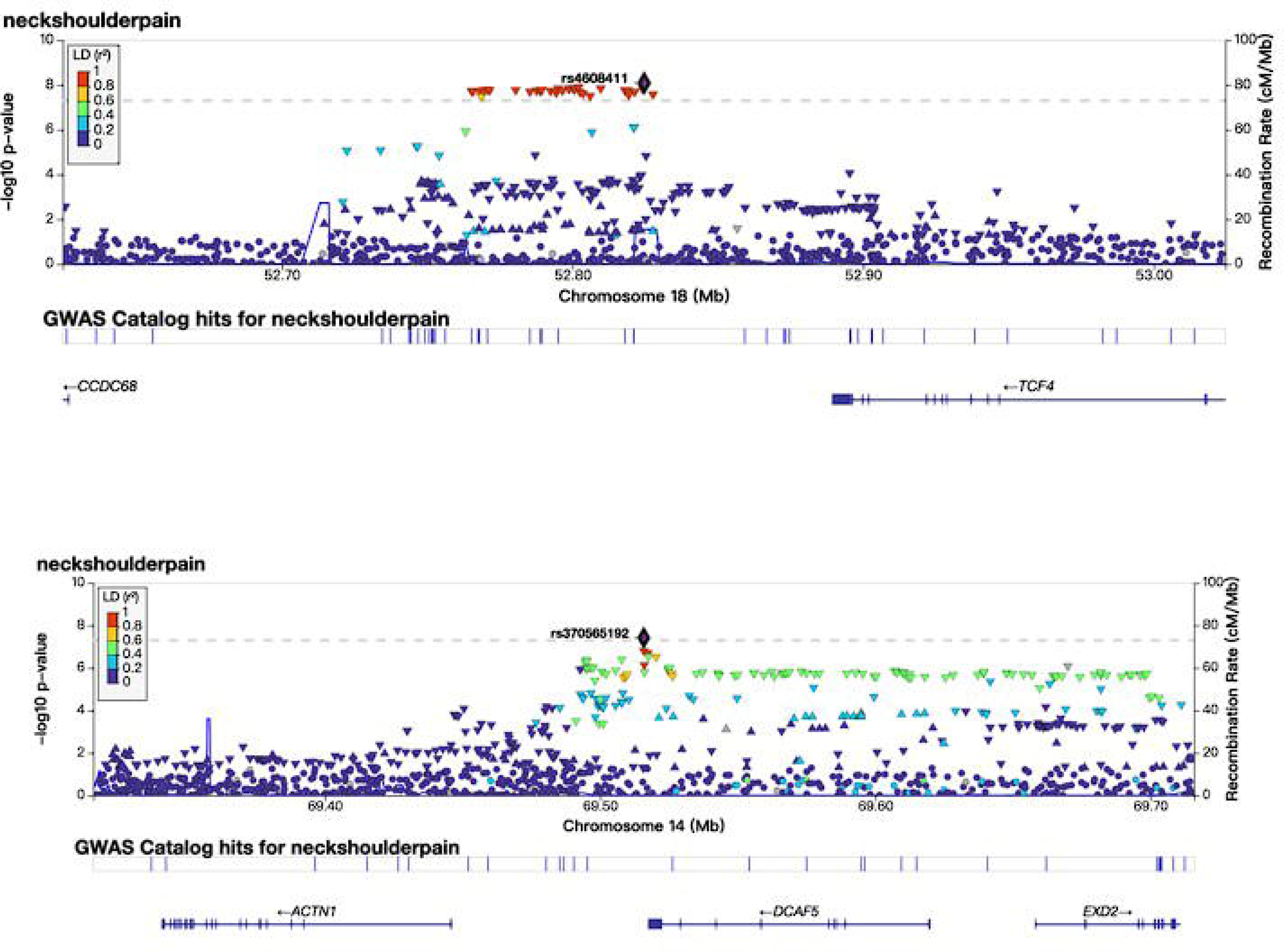
The regional plots of locus in the TCF4 and DCAF5 regions

In the secondary GWAS analyses, which were stratified by sex, the male-specific GWAS identified a single locus of genome-wide significance associated with neck or shoulder pain. This locus, distinct from those found in the primary GWAS, was located in the *SLC24A3* gene on chromosome 20, with a lowest *p* value of 6.52 x 10^-9^ for rs16980973. The female-specific GWAS revealed two significant loci associated with neck or shoulder pain, one of which differed from the primary GWAS findings. This novel locus was identified near the *LINC02770* gene on chromosome 1, with a *p* value of 3.57 x 10^-8^ for rs5779595. Detailed information on these findings is available in Table 3, and the corresponding Manhattan plots can be found in Figure 1.

**Table 3.**
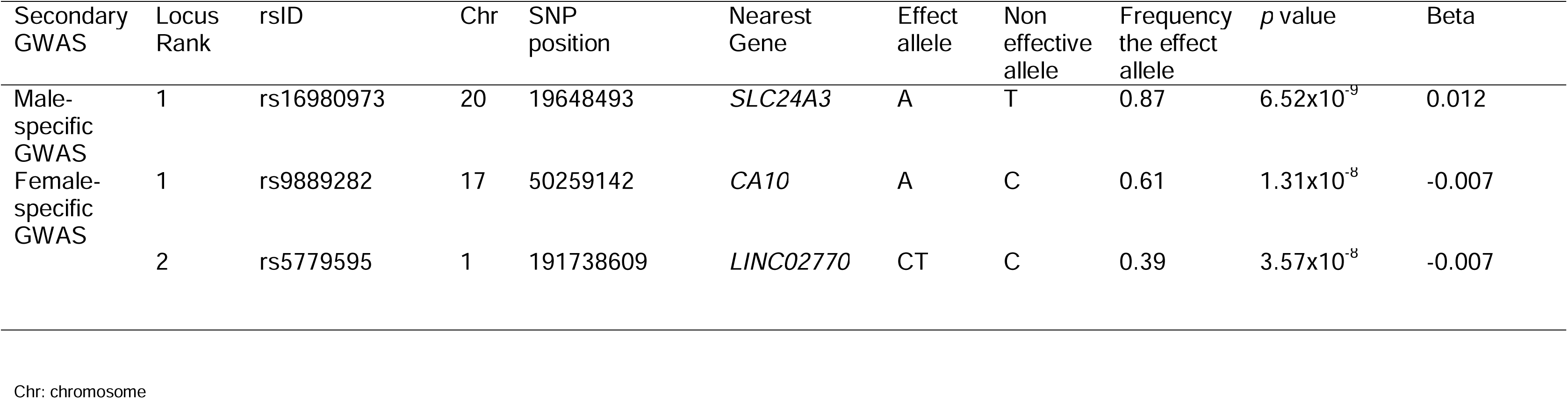
The top SNPs within 3 loci identified by the male-specific GWAS and female-specific GWAS on neck or shoulder pain.

### Gene, gene-set and tissue expression analysis by FUMA

In our gene analysis of primary GWAS, *FOXP2* displayed the most significant association, with a *p* value of 1.69 x 10^-9^. A total of six genes, namely *PABPC4, TCTA, MICB, FOXP2, IST1* and *SLC44A2*, were found to be associated with neck or shoulder pain, each with a *p* value less than 2.60 x 10^-6^ (0.05/19203). The detailed results of this analysis can be found in Supplementary Table 2.

In the gene-set analysis using MEGMA, a comprehensive examination of 15,485 gene sets was conducted. Of these, one gene set, specifically ‘GOCC PRESYNAPTIC ENDOCYTIC ZONE’, was identified as significantly associated (*p* = 2.95 x 10^-7^). The top ten gene sets from this analysis have been detailed in Supplementary Table 3.

In the tissue expression analysis, the broader categories of ‘Brain’ and ‘Pituitary’ showed statistical significance among 30 general tissue types across various organs. Notably, eight brain-associated tissues, including ‘Brain Cortex,’ ‘Brain Frontal Cortex BA9’, ‘Brain Anterior Cingulate Cortex BA24’, ‘Brain Caudate Basal Ganglia’, ‘Brain Nucleus Accumbens Basal Ganglia’, ‘Brain Hypothalamus’, ‘Brain Hippocampus’ and ‘Brain Amygdala’ exhibited significant associations (*p* < 0.001) out of 53 specific tissue types from different organs. Further details and visualizations of these findings can be found in Figures 4 and 5.

**Fig. 4.**
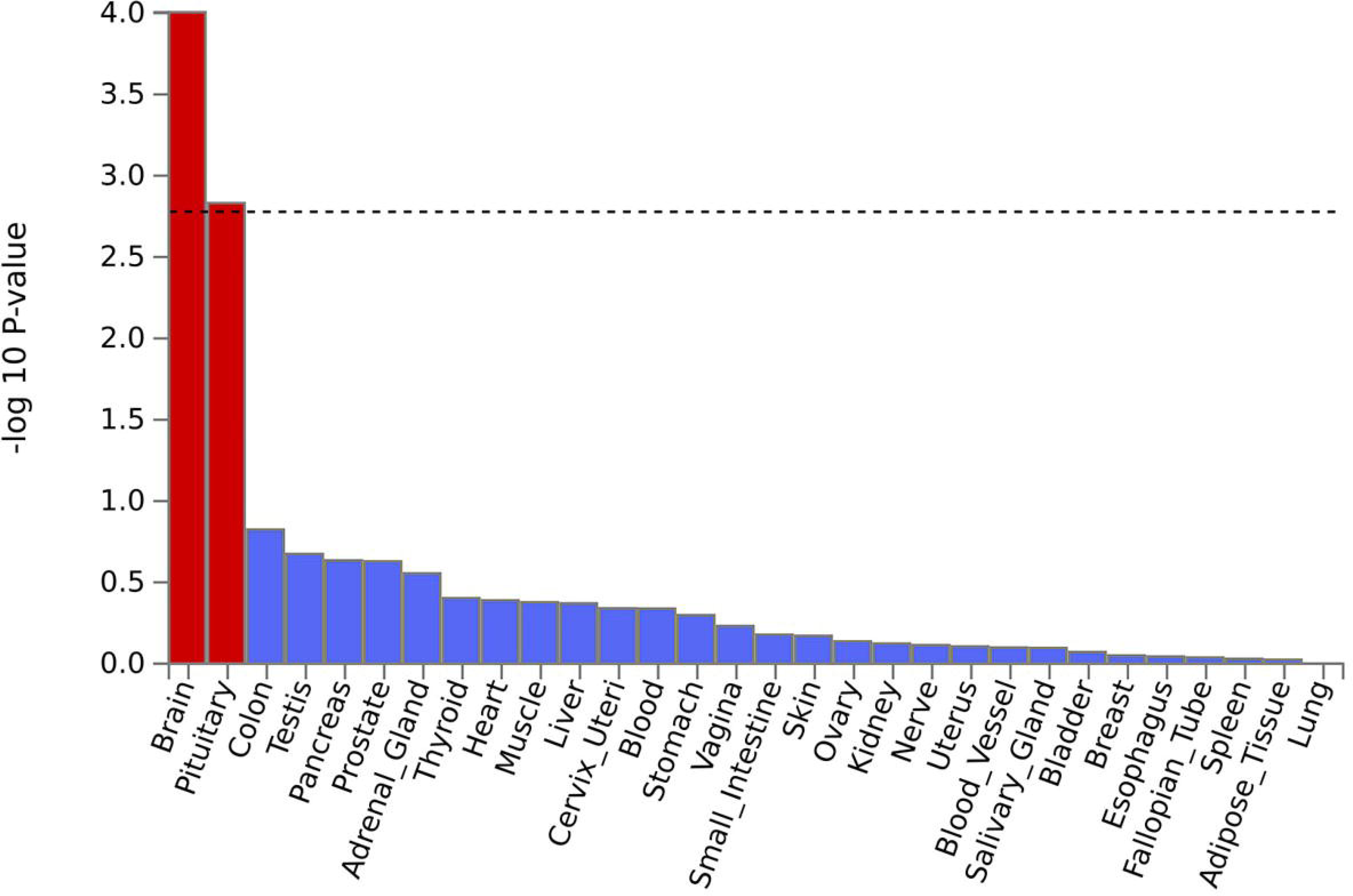
Tissue expression results on 30 specific tissue types by GTEx in the FUMA The dashed line shows the cut-off *p* value for significance with Bonferroni adjustment for multiple hypothesis testing

**Fig. 5.**
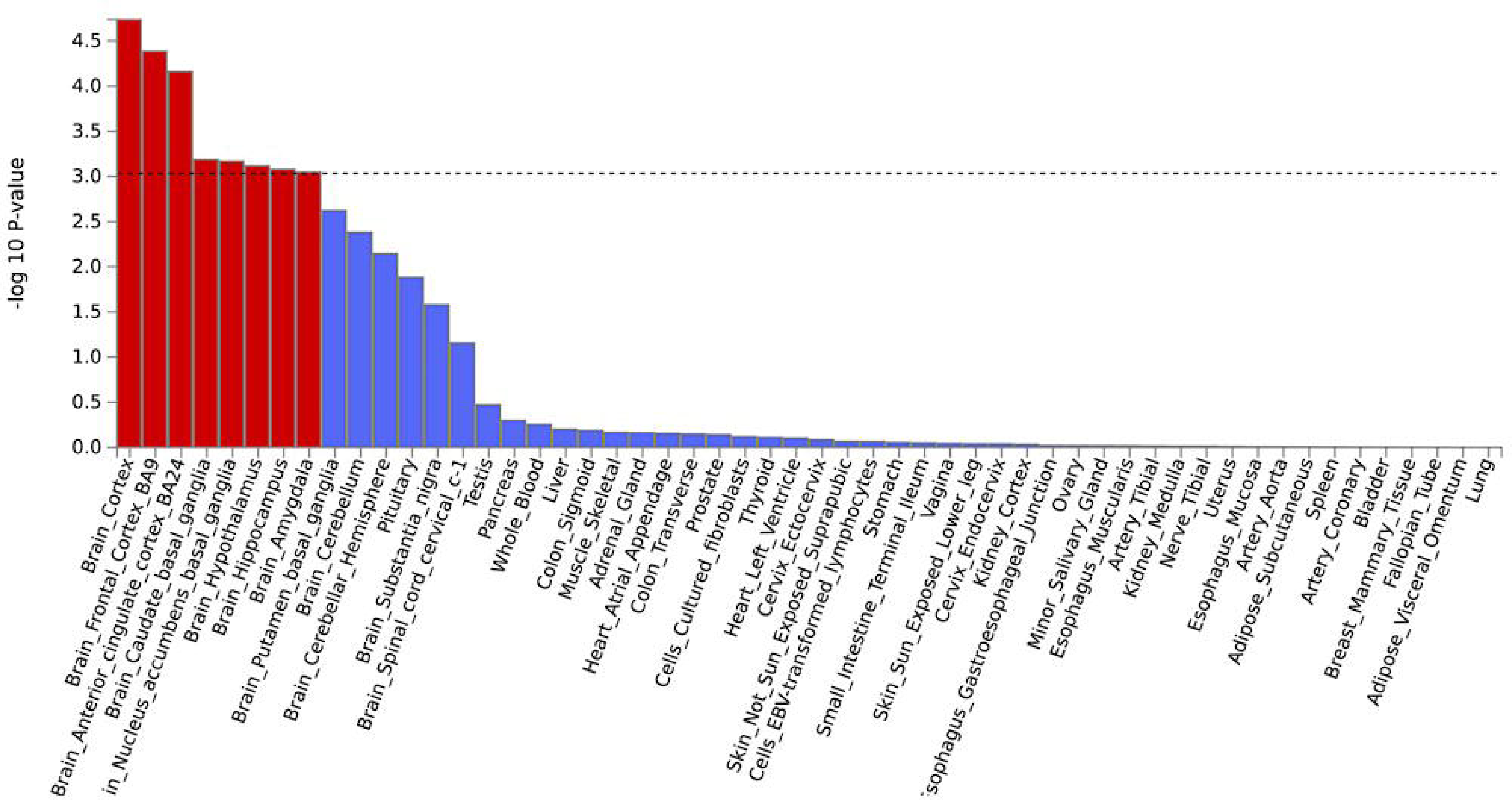
Tissue expression results on 53 specific tissue types by GTEx in the FUMA The dashed line shows the cut-off *p* value for significance with Bonferroni adjustment for multiple hypothesis testing

### Genetic correlation analysis

In our analysis of the genetic correlations between neck or shoulder pain and other phenotypes, several significant associations were identified. Neck or shoulder pain showed substantial genetic correlations with some other pain phenotypes, including multisite chronic pain (*r_g_* = 0.89, *p* = 0), back pain (*r_g_* = 0.82, *p* = 1.06 x 10^-175^), and hip pain (*r_g_* = 0.77, *p* = 9.26 x 10^-91^). Additionally, significant positive genetic correlations were observed with neck or shoulder pain in relation to various medical conditions and health outcomes, including painful gums (*r_g_* = 0.72, *p* = 2.81 x 10^-23^), usage of amitriptyline (*r_g_* = 0.74, *p* = 1.26 x 10^-16^), and usage of co-codamol (*r_g_* = 0.76, *p* = 6.09 x 10^-38^). These results are comprehensively documented in Supplementary Table 4 and depicted in Figure 6. The genetic correlation for neck or shoulder pain between males and females was also calculated (*r_g_* = 0.79, *p* = 2.67 x 10^-41^), potentially providing extra evidence for the observed genetic differences in the sex-stratified secondary GWAS.

**Fig.6.**
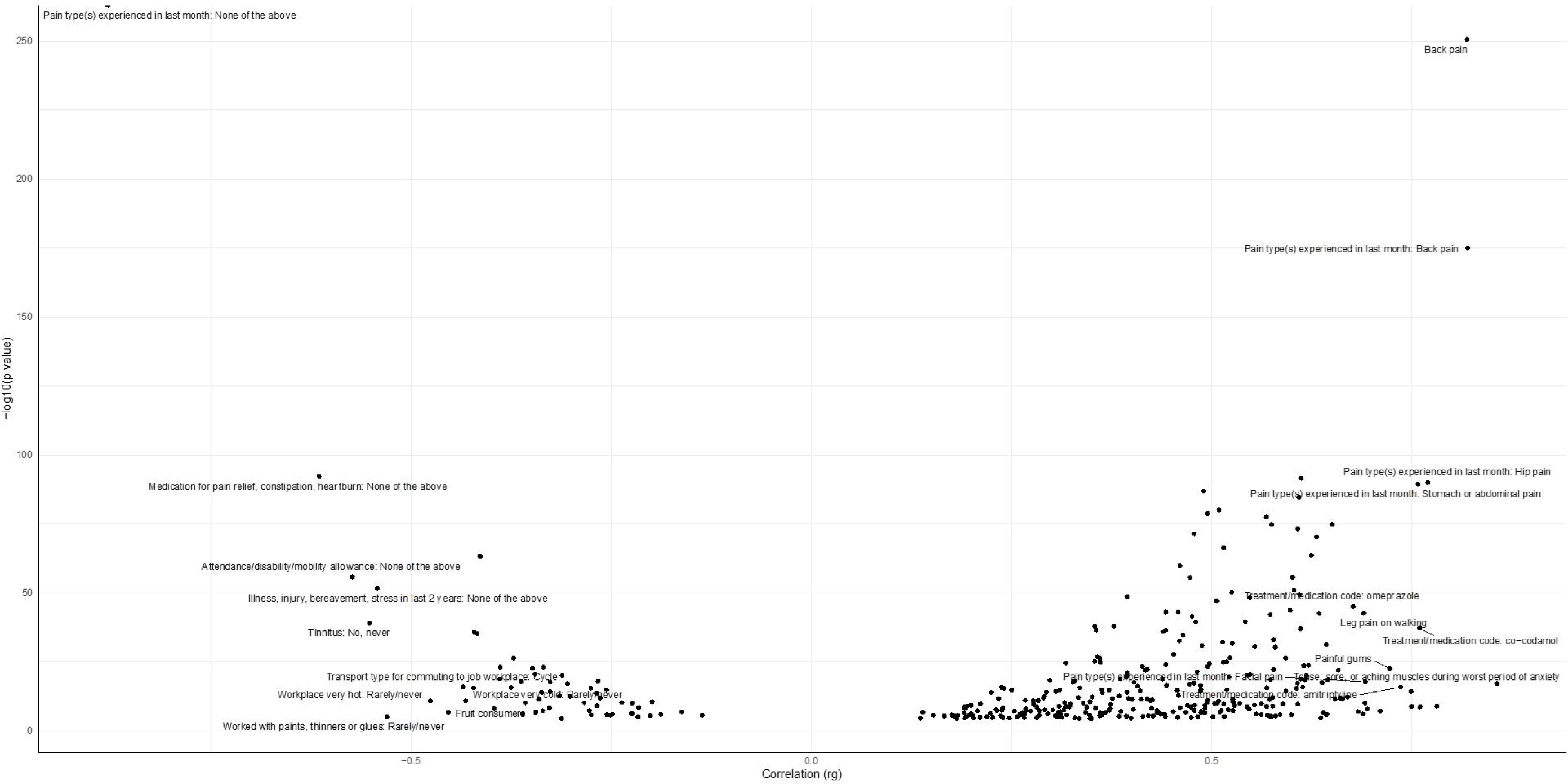
Genetic correlation results for pain intensity using LDSC on CTG-VL. For a full list, see Supplementary Table 4

### PheWAS

PheWAS was conducted using the GWAS ATLAS platform to investigate phenotypes associated with independent significantly associated SNPs (rs9889282, rs34291892, rs13135092, rs4608411, rs370565192, rs12951067), and significantly associated genes (*SLC39A8*, *FOXP2*, *DCAF5*, *CA10*, and *TCF4*). rs9889282 was significantly associated with the self-rated health trait (*p* = 3.82 x 10^-7^), while rs34291892 demonstrated a significant association with educational attainment (*p* = 1.70 x 10^-6^). Both rs13135092 and rs4608411 were associated with schizophrenia (*p* = 3.60 x 10^-12^ and *p* = 7.42 x 10^-5^, respectively). rs370565192 was associated with the Intelligence trait (*p* = 3.87 x 10^-5^), and rs12951067 was associated with the frequency of tiredness/lethargy in the last 2 weeks (*p* = 4.25 x 10^-5^). The genes *SLC39A8*, *FOXP2*, and *DCAF5* demonstrated associations with many traits following Bonferroni correction, covering a range of domains including ophthalmological, cellular, psychiatric, immunological, cardiovascular, and metabolic areas. The *CA10* and *TCF4* genes exhibited a strong association with psychiatric traits, specifically ‘morningness’ (*p* = 5.42 x 10^-11^ for *CA10*) and schizophrenia (*p* = 3.64 x 10^-20^ for *TCF4*). All significant traits that passed Bonferroni correction, are listed in Supplementary Table 5. The plots of phenotypes associated with SNPs and genes are visualized in Supplementary Figures 3 and 4, respectively.

## Discussion

These three GWAS on neck or shoulder pain utilizing the UK Biobank dataset identified several notable genetic loci associated with the phenotype, including five loci (two novel) in the primary analysis, two loci in females and one locus in males (Table 2 and 3).

In the primary GWAS, the locus near *CA10* on chromosome 17 was the most significantly associated with neck or shoulder pain, with a top SNP of rs9889282 (*p* = 2.63 x 10^-12^). *CA10* encodes a protein from the zinc metalloenzyme family called carbonic anhydrase, involved in bone resorption and bone mineral solubilization (Okamoto et al. 2001; Pastorekova et al. 2004). The protein is also believed to be involved in the development of the brain and the central nervous system (https://www.ncbi.nlm.nih.gov/gene/105371829#gene-expression). The strong interconnections between carbonic anhydrase activity and neuropathic pain have been proved by Asiedu et al.(2010, 2014). Da Vitoria Lobo et al. (2022) concluded through experiments that inhibition of carbonic anhydrase activity by intraperitoneal injection of acetazolamide decreased neuropathic pain. For *CA10,* one study suggested that it has at least two global functions that do not interact (Sterky et al. 2017). First, overexpression of *CA10* results in increased surface levels, altered posttranslational modification patterns, and total levels of neuroxin proteins (Sterky et al. 2017). Secondly, *CA10* may play the role of adaptors, facilitating neurexins’ indirect connections with unidentified postsynaptic target molecules, and thus mediating the development of new transsynaptic complexes (Sterky et al. 2017). Evidence suggests that the mechanism by which regulatory proteins interact with neurexins-neuroligins to jointly form intersynaptic bridges plays a role in pathological pain (Li et al. 2022). As there are not many studies on *CA10* in humans, future studies should focus on elucidating the mechanism of *CA10* action on neck or shoulder pain.

The second most significantly associated locus was identified in the *FOXP2* gene located on chromosome 7, with a top SNP of rs34291892 (*p* = 1.69 x 10^-9^). *FOXP2* is one member of the forkhead family of transcription factors and is located on human chromosome 7q31, which encodes a 715-amino acid protein (Enard et al. 2002). It comprises a glutamine-rich area made up of two neighboring polyglutamine tracts encoded by CAG and CAA repeat mixes, which are known to have high mutation rates (Enard et al. 2002). The *FOXP2* gene may play a significant role in the establishment of a putative frontostriatal network, which is associated with the function of learning, planning, and execution in the speech motor sequences, similar to that engaged in other types of motor abilities (Liégeois et al. 2003). In the avian striatum, *FOXP2* expression is controlled by singing, suggesting the prospect that developmental verbal dyspraxia resulting from human *FOXP2* mutation may largely represent a loss in ongoing brain signaling, rather than developmental miswiring (Deshpande and Lints 2013). Alternatively, genes *DLX5* and *SYT4*, which are affected by *FOXP2*, are essential for brain development and function (Meng et al. 2020). In a previous GWAS analysis, *FOXP2* has been confirmed to be associated with neck or shoulder pain, and multi-site chronic pain (Johnston et al. 2019; Meng et al. 2020). In addition, *FOXP2* has been found to be associated with the development of attention deficit hyperactivity disorder and psychological disorders (Wendt et al. 2018; Meyer et al. 2023). The identified brain regions, including the hypothalamus, are known to play crucial roles in central neuroendocrine and hormone secretion (Markakis 2002). Studies of post-mortem brain microscopy have shown a link between the hypothalamus and mental illness(Bernstein et al. 2019). This connection raises questions about the psychologicalaspects influencing the development and persistence of neck or shoulder pain, which is consistent with the function.

The third notable association was identified in the *SLC39A8* gene on chromosome 4. *SLC39A8* encodes transporter zinc- and iron-related protein eight (ZIP8), which carry essential-metal divalent cations including zinc, copper, iron, and manganese (Gálvez-Peralta et al. 2012). In a previous study, foetal and neonatal mice with *SLC39A8* homozygote show reduced numbers of hematopoietic islands in the hematocrit, serum iron, red cell count, yolk sac and liver, low hemoglobin levels, and total iron-binding capacity—all consistent with the presence of severe anemia (Nebert and Liu 2019). GWAS subsequently revealed that human *SLC39A8*-deficient variants exhibit distinct multiorientation defects associated with clinical diseases in almost all organs including many developmental and congenital diseases, musculoskeletal system, central nervous system, coagulation system, cardiovascular system and immune system (Nebert and Liu 2019). In the details of the central nervous system, target validation tests demonstrated that microRNA 488 can target validation tests demonstrated that microRNA 488 can target *SLC39A8* mRNA, and inhibiting *SLC39A8* expression in the osteoarthritis animal model reduced cartilage deterioration (Song et al. 2013). A subsequent paper has suggested that the increased expression of *SLC39A8* is associated with increased intracellular Zn levels in diseased chondrocytes among all Zn transporters in human and mouse cartilage affected by osteoarthritis (Kim et al. 2014). Musculoskeletal conditions, including osteoarthritis, can contribute to pain and discomfort in the neck and shoulder regions, making *SLC39A8* a potential player in these processes. The positive correlation between *SLC39A8* and osteoarthritis in the experiment with human and mouse embryos corroborates this conjecture. It is worth noting that this locus near *SLC39A8* has already been found to be associated with chronic neck or shoulder pain in a GWAS analysis of chronic musculoskeletal pain (Tsepilov et al. 2020).

The two novel loci are near *TCF4* on chromosome 18 with a top SNP of rs4608411 (*p* = 8.20 x 10^-9^) and in *DCAF5* on chromosome 14 with a top SNP of rs370565192 (*p* = 3.80 x 10^-8^), separately. The discovery of these two new loci represents a significant further step forward in unraveling the complex genetic architecture of neck or shoulder pain.

*TCF4* encodes transcription factor 4, a fundamental helix-loop-helix transcription factor. The encoded protein is a member of the ubiquitous E-protein family, causes severe CNS and autonomous nervous system dysfunction when mutated in humans (Amiel et al. 2007). Rare variants of *TCF4* including nonsense, missense, or microdeletion mutations may cause haploidy deficiency, which leads to a syndromic encephalopathy called Pitt-Hopkins syndrome (Brockschmidt et al. 2007; de Pontual et al. 2009; Forrest et al. 2012; Sweatt 2013; Li et al. 2019). In certain circumstances, it is harmful of the large amount of mutations missense mutations or create shortened proteins to the bHLH-binding domain crucial for the functional transcription activity of *TCF4* (Sepp et al. 2012; Sweatt 2013; Li et al. 2019). Mutations in *TCF4* have also been linked to a number of other psychiatric disorders including autism, bipolar disorder and schizophrenia (Stefansson et al. 2009; Steinberg et al. 2011; Forrest et al. 2014; Hamdan et al. 2014). Meanwhile, *TCF4* shows expression in the adult brain, skeletal muscle, lung, and heart, confirming the disease’s postnatal growth retardation and microcephaly (Amiel et al. 2007).

The *DCAF5* gene is one part of the ubiquitin ligase complex *DDB1-CUL4*, and the *DCAF5* gene’s brain expression results in *DDB1*-and-*CUL4* related factor 5 (Angers et al. 2006; Lee and Zhou 2007; Śmigiel et al. 2020). *CUL4B* makes up the majority of this complex, and mutations in it can result in X-linked intellectual impairment (Tarpey et al. 2007). Likely haploinsufficiency of genes including *DCAF5* deletion are related to congenital heart defects, short fingers, and mild intellectual disability (Oehl-Jaschkowitz et al. 2014). It has been proposed that dysfunction of its binding partner *DCAF5* caused by de novo mutations may have similar consequences for developmental function (Śmigiel et al. 2020). *DCAF5* carries rare segregating variants in at least two independent families with bipolar disorder, which are hypothesized to be associated with psychiatric disorders (Forstner et al. 2020). In addition, a GWAS related to lung function identified rare functional coding variants in *DCAF5* that prevented rapid lung function decline (Mathias et al. 2012).

In the sex-stratified GWAS analyses, male-specific GWAS revealed a locus n *SLC24A3* on chromosome 20, with a top SNP of rs16980973 (*p* = 6.52 x 10^-9^). *SLC24A3* encodes NCKX3, a potassium-dependent Na+/K+/Ca2+ exchanger that regulates intracellular calcium homeostasis (Citterio et al. 2011). Ca^2+^ has an important function in modulating vascular tone, which contributes significantly to the regulation of systemic blood pressure (Karaki et al. 1997; Blaustein and Lederer 1999). Because there is not sufficient evidence to clarify the uniqueness of male-specific locus associated with neck or shoulder pain, more research should focus on the differences in neck or shoulder pain between males and females.

In female-specific GWAS analysis, the most significant variant was identified near *CA10* same as the primary GWAS. The other significant variant is near the *LINC02770* gene on chromosome 1 with the top SNP rs5779595 (*p* = 3.57 x 10^-8^). In a genome-wide association study investigating associations with obstructive sleep apnea, *LINC02770* has been identified as being linked to this sleep disorder (Mohit et al. 2022). *LINC02770* is also thought to be associated with insomnia and post-traumatic stress disorder (https://www.genecards.org/cgi-bin/carddisp.pl?gene=LINC02770). These findings align with clinical observations where individuals experiencing chronic neck or shoulder pain often report associated symptoms such as headaches, insomnia and mental illness including depression (Shahidi et al. 2015; Chu and Lee 2022). Although the specific genetic mechanism of *LINC02770* has not been explored, it introduces a novel aspect to the understanding of genetic factors contributing to pain susceptibility in females. The function of *LINC02770* and its potential involvement in pain modulation warrant further investigation to elucidate the intricate mechanisms. The genetic correlation for neck or shoulder pain between males and females was also calculated (*r_g_* = 0.79), which suggests that the genetic mechanisms of neck or shoulder pain are not the exactly same between males and females. The reason for the difference is likely due to the important role of gender differences in the heterogeneity of pain, especially neuropathic pain (Gao et al. 2024; Smith 2024). These sex-specific genetic associations underscore the importance of considering biological differences between males and females in studies related to pain genetics. Tailoring interventions according to the identified genetic factors could lead to more effective and targeted pain management strategies, ultimately improving the quality of life for individuals suffering from these conditions.

Compared to the previous GWAS on neck or shoulder pain by Meng et al. (2020), this study discovered two new loci. In this study, the SNP-based heritability for neck or shoulder pain was determined to be 0.05, which is smaller than the previously estimate (0.115) by Meng et al. (2020). The likely reason is the different definition of controls, which leads to a larger number of people in the control group. The scope of the control group in this study is larger than that of the former. Since the scope of the case group is unchanged, a larger number of controls is likely to lead to a decrease in the SNP heritability of the GWAS result.

In the tissue expression analysis, neck or shoulder pain showed a strong correlation with the brain and pituitary categories. The identified brain tissues revealed key signaling pathways related to pain perception (BA24, hypothalamus, amygdala), sensory processing (amygdala, cortex, BA9, basal ganglia), and central neuroendocrine (hypothalamus) (Hudson 2000; Peyron et al. 2000; Markakis 2002; Kerestes et al. 2012; Lanciego et al. 2012). These findings align with clinical observations where individuals experiencing chronic neck or shoulder pain often report associated symptoms such as headaches, fatigue, and alterations in mood (Shahidi et al. 2015; Chu and Lee 2022). The identified brain regions, including the cerebral cortex and amygdala, are known to play crucial roles in emotional processing and stress response (Šimić et al. 2021). This connection raises questions about the psychosocial aspects influencing the development and persistence of neck and shoulder pain. Moreover, tissue specific expression in the pituitary gland displayed a significant association with neck and shoulder pain. This finding suggests a potential involvement of hormonal regulation in the manifestation of pain in these regions, and could be at least part of the explanation for the sex differences. The intricate network between the brain, pituitary gland, and chronic pain in the neck or shoulder highlights the need for a comprehensive understanding of both neurological and endocrine factors contributing to this phenomenon.

The genetic correlations between neck or shoulder pain and several other pain phenotypes suggest that these disorders share common genetic determinants. The three most highly correlated phenotypes of neck or shoulder pain were multisite chronic pain (*r_g_* = 0.89), back pain (*r_g_* = 0.82), and hip pain (*r_g_* = 0.77). In addition, a correlation emerged between neck or shoulder pain and gum pain (*r_g_* = 0.74), highlighting the interconnectedness of these symptoms. Understanding these correlations could provide valuable insights into potential shared pathways or mechanisms underlying these health outcomes. Neck or shoulder pain was observed to be associated with various medical conditions and health outcomes. Moving beyond genetic correlations, the associations with specific medications add another layer of complexity. The correlation between neck or shoulder pain and receipt of amitriptyline (*r_g_* = 0.74) and co-codamol (*r_g_* = 0.76) suggests that biological links exist between the neck or shoulder pain and each phenotype as amitriptyline is an anti-depression drug although not used as a first-line treatment for depression due to considerable side effects and co-codamol is a pain-relief medicine. In the PheWAS, the results demonstrate that the top SNPs and corresponding genes are primarily highly associated with numerous psychiatric traits. This may provide further evidence that psychiatric factors could be an important contributor to neck or shoulder pain and/or vice versa. Meanwhile, these newly identified SNPs and genes are likely to play a role in the biological mechanism of neck or shoulder pain, which will have a suggestive effect on the development of targeted therapies (such as carbonic anhydrase inhibitors) for the pain. The shared genetic determinants, correlations with other pain phenotypes, and connections to various medical conditions and medications underscore the need for a holistic approach in understanding and addressing neck or shoulder pain.

It is important to note that cases and controls in the UK Biobank are self-reported and may be subject to several biases such as social expectations, recall periods, sampling methods or selective recall. Therefore, these potential effects need to be carefully considered when interpreting the findings. A potential limitation is that the control group contains persons with pain at other body sites. This may have an impact on the GWAS results. Genetic correlation studies revealed that many pain phenotypes are strongly genetically related. For example, as identified above (REF) there is a particularly strong genetic correlation between back pain and neck or shoulder pain. As a sensitivity analysis we repeated the above GWAS after excluding people who reported back pain in the control group. This re-analysis led to similar results in the genetic associations identified (Supplementary Figure 5 and Supplementary Table 6). Additionally, we described cases and controls with neck or shoulder pain based on responses to specific questions from UK Biobank participants. However, this question was not designed to cover more detailed information about the severity, frequency, and exact location of the pain. Therefore, the phenotypic definitions we derived from these responses should be viewed as generalized. Since the sample size in this study was very large (about 450,000), it has sufficient study power to detect contributing genetic variants. We noticed that the *p* values of some loci associations were just smaller than the GWAS significance, therefore it is likely that the results can only be replicated by cohorts of a similar size as the UK Biobank, or larger. Neck or shoulder pain phenotypes were also not recorded by many other large publicly available genetic cohorts such as FinnGen (Kurki et al. 2023), and we therefore did not conduct replication analysis.

## Conclusion

In summary, our primary GWAS identified five genetic loci including two novel ones associated with neck or shoulder pain among all people with neck or shoulder pain. Our secondary GWAS identified a single novel genetic locus associated with neck or shoulder pain among males and two genetic loci (including one novel) associated with neck or shoulder pain among females. These discoveries not only expand our understanding of the genetic underpinnings of neck or shoulder pain but also emphasize the importance of considering sex-specific influences.

## Data Availability and Acknowledgement

This study adheres to all ethical guidelines and data protection protocols of the UK Biobank. The current study was conducted under approved UK Biobank data application number 89386. The summary statistics of the UK Biobank results on neck or shoulder pain can be accessed upon publication. Any other data relevant to the study that are not included in the article or its supplementary materials are available from the authors upon reasonable request.

## Author information

### Authors’ Contributions

All authors contributed to the analysis plan. YT and QP performed the UK Biobank GWAS analysis and drafted the paper. TC contributed to data formatting. ZHL, MH, TD, LAC and BHS provided comments to the paper. WM organised the project and provided comments.

### Corresponding authors

Correspondence to Weihua Meng.

### Consent to Publish

All authors provide consent for publication.

### Ethical Approval

This study was approved by the Ethics Committee of the University of Nottingham Ningbo China.

