## Supplementary figures and images for "A Genome-wide Association Study Identifies Novel Genetic Variants Associated with Neck or Shoulder Pain in the UK Biobank (N = 441,757)"

### Supplementary Figures 1

#### Slide 1
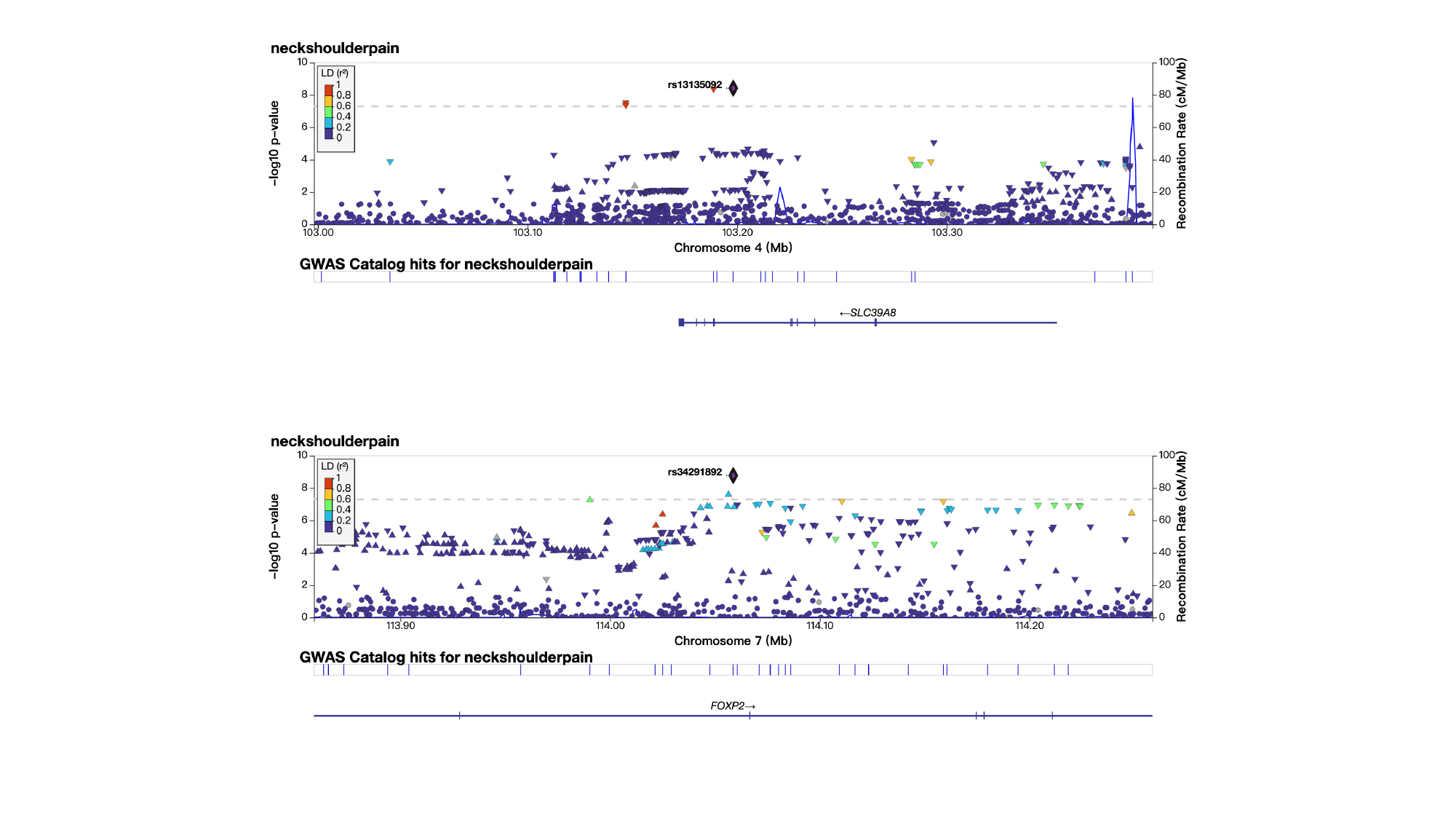

### Supplementary Figures 2

#### Slide 1
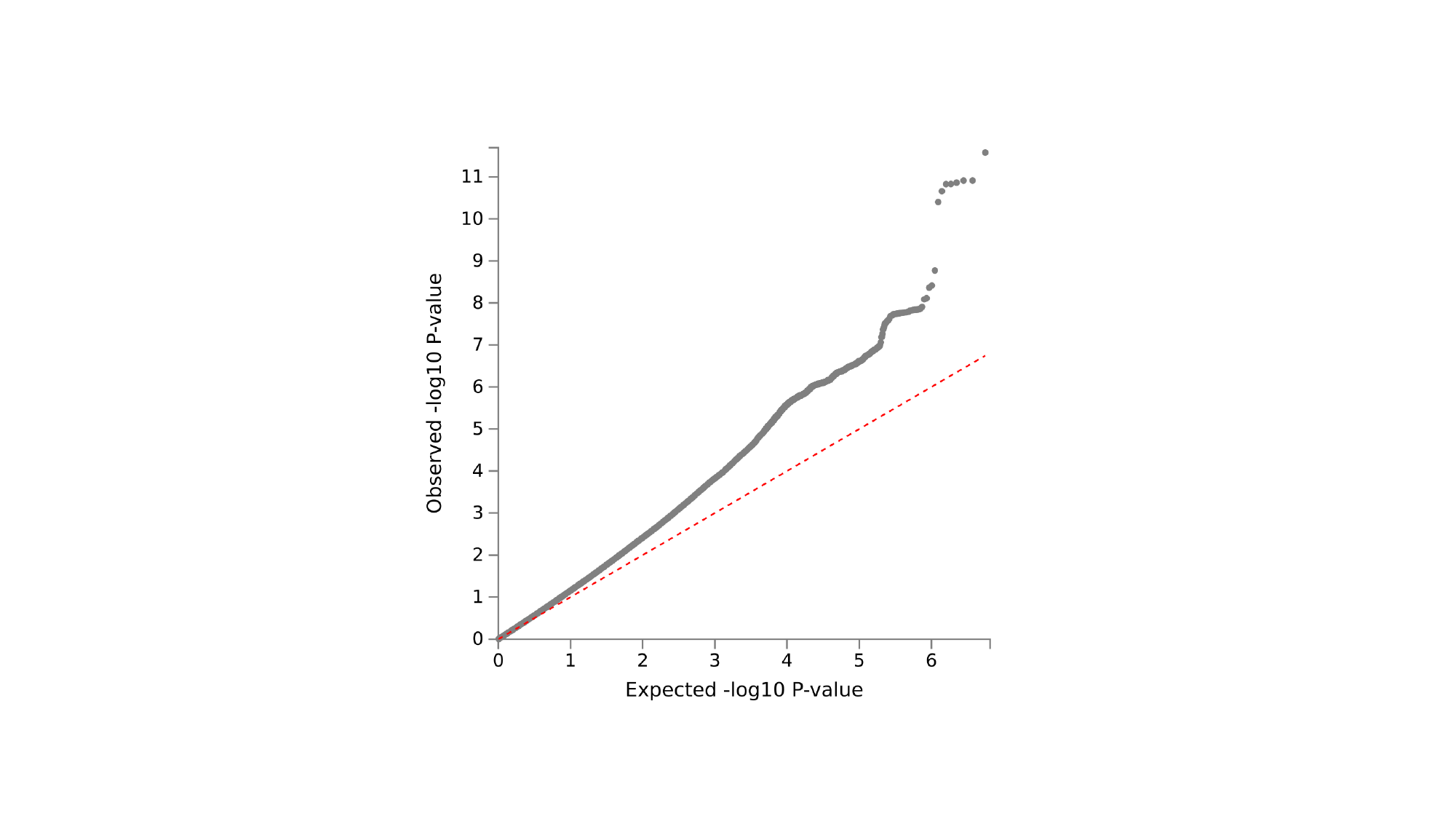

### Supplementary Figures 5

#### Slide 1
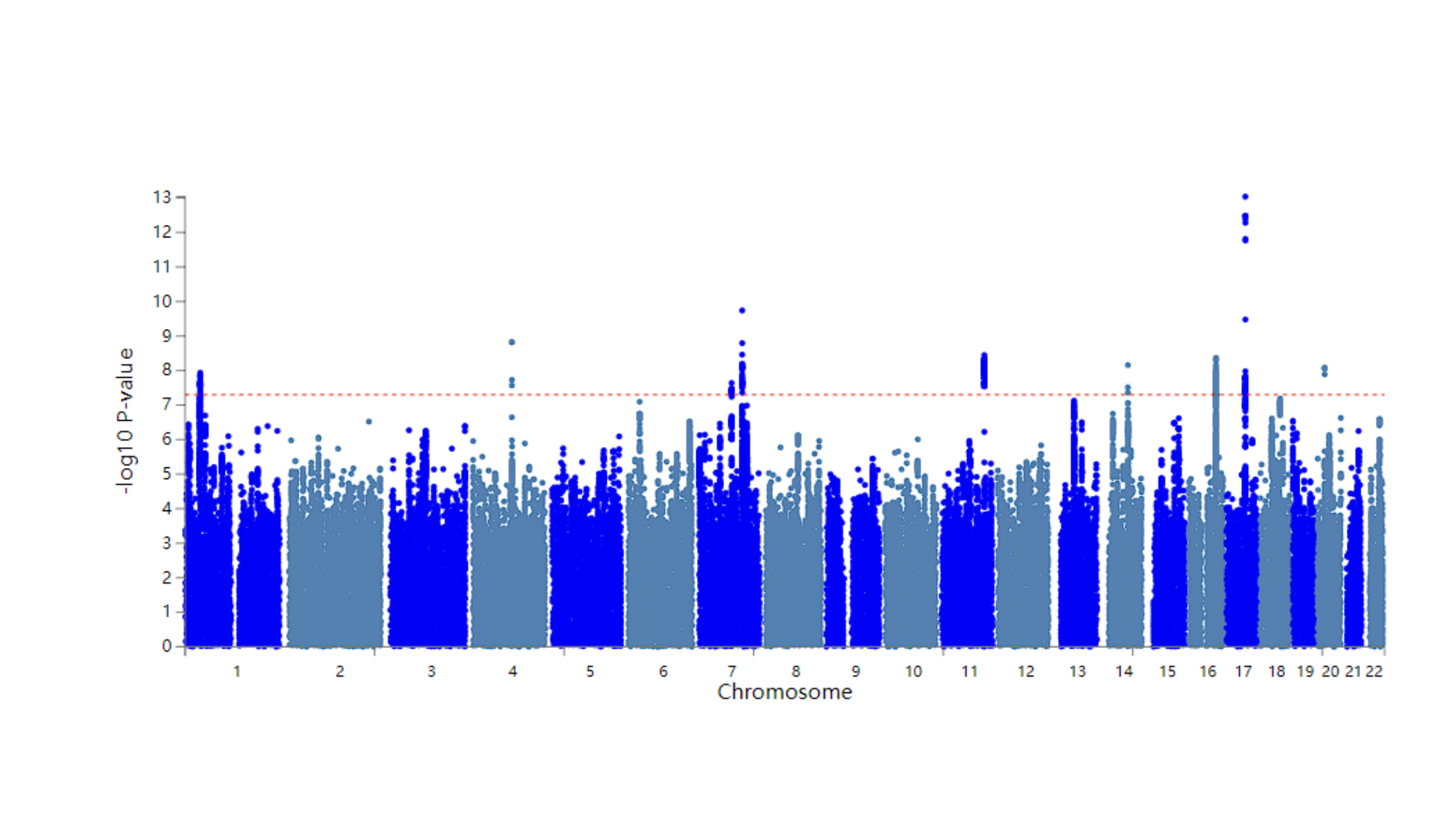
