## Supplementary Figures 3 for "A Genome-wide Association Study Identifies Novel Genetic Variants Associated with Neck or Shoulder Pain in the UK Biobank (N = 441,757)"

### Slide 1
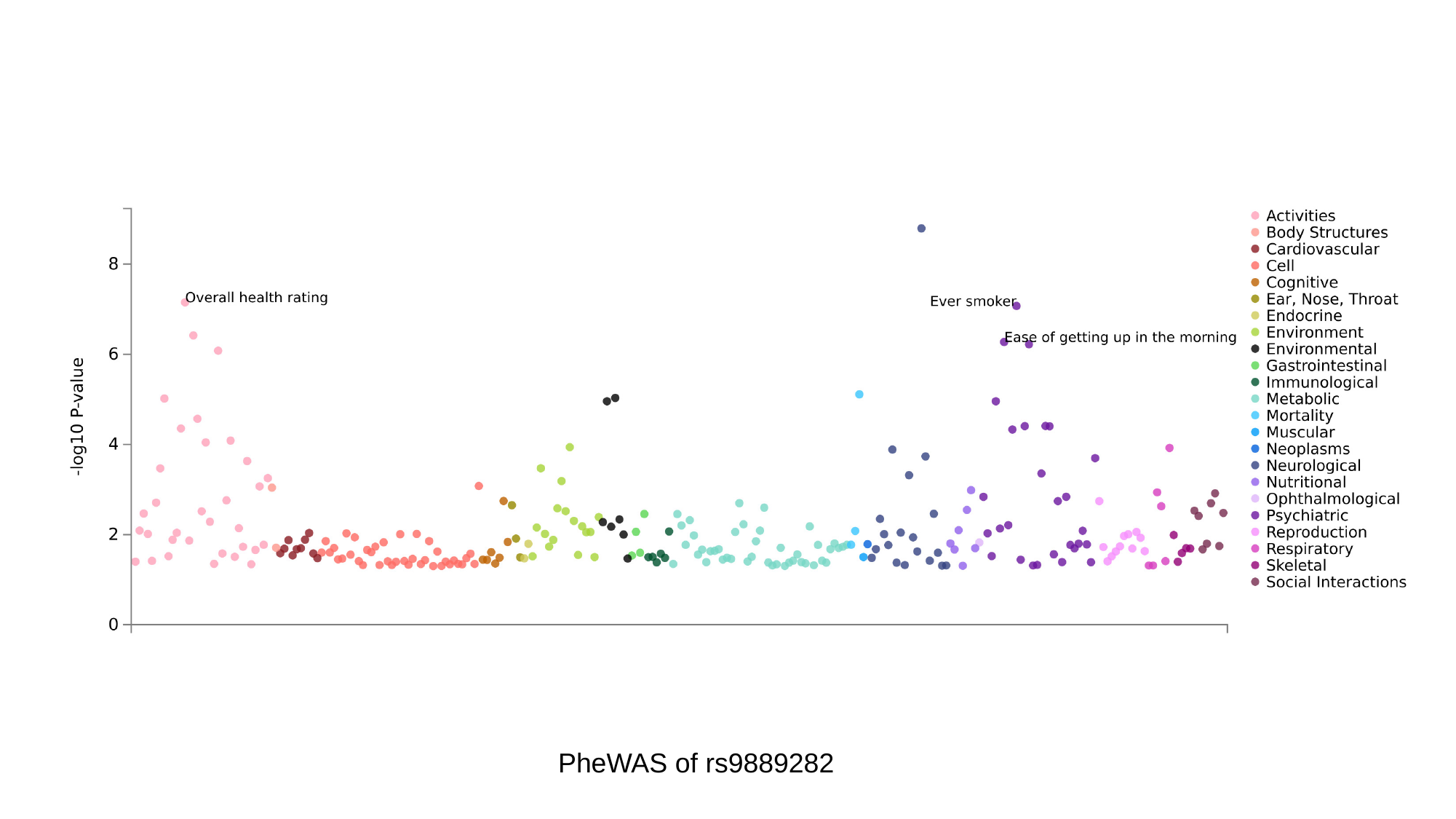

PheWAS of rs9889282

### Slide 2
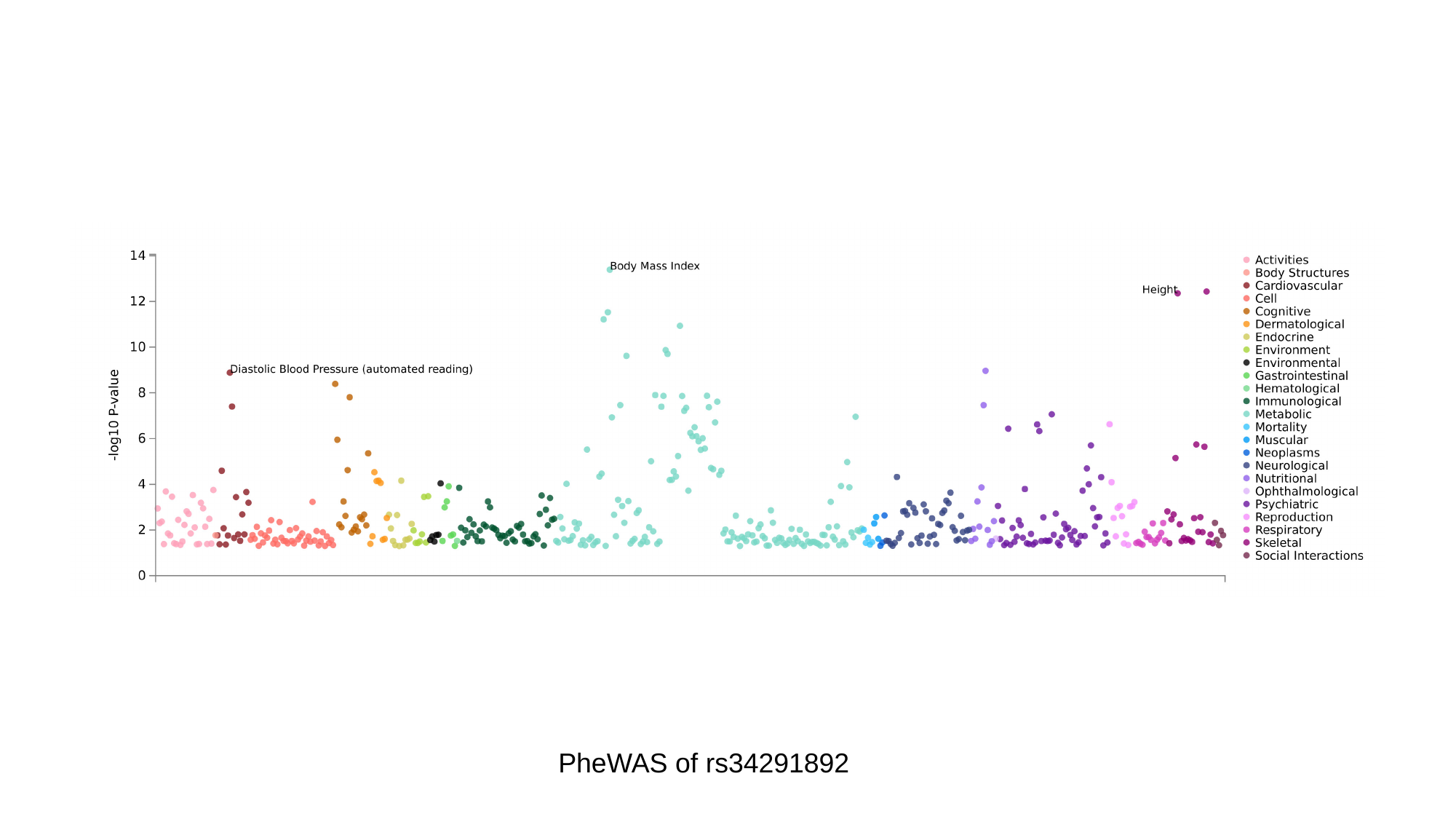

PheWAS of rs34291892

### Slide 3
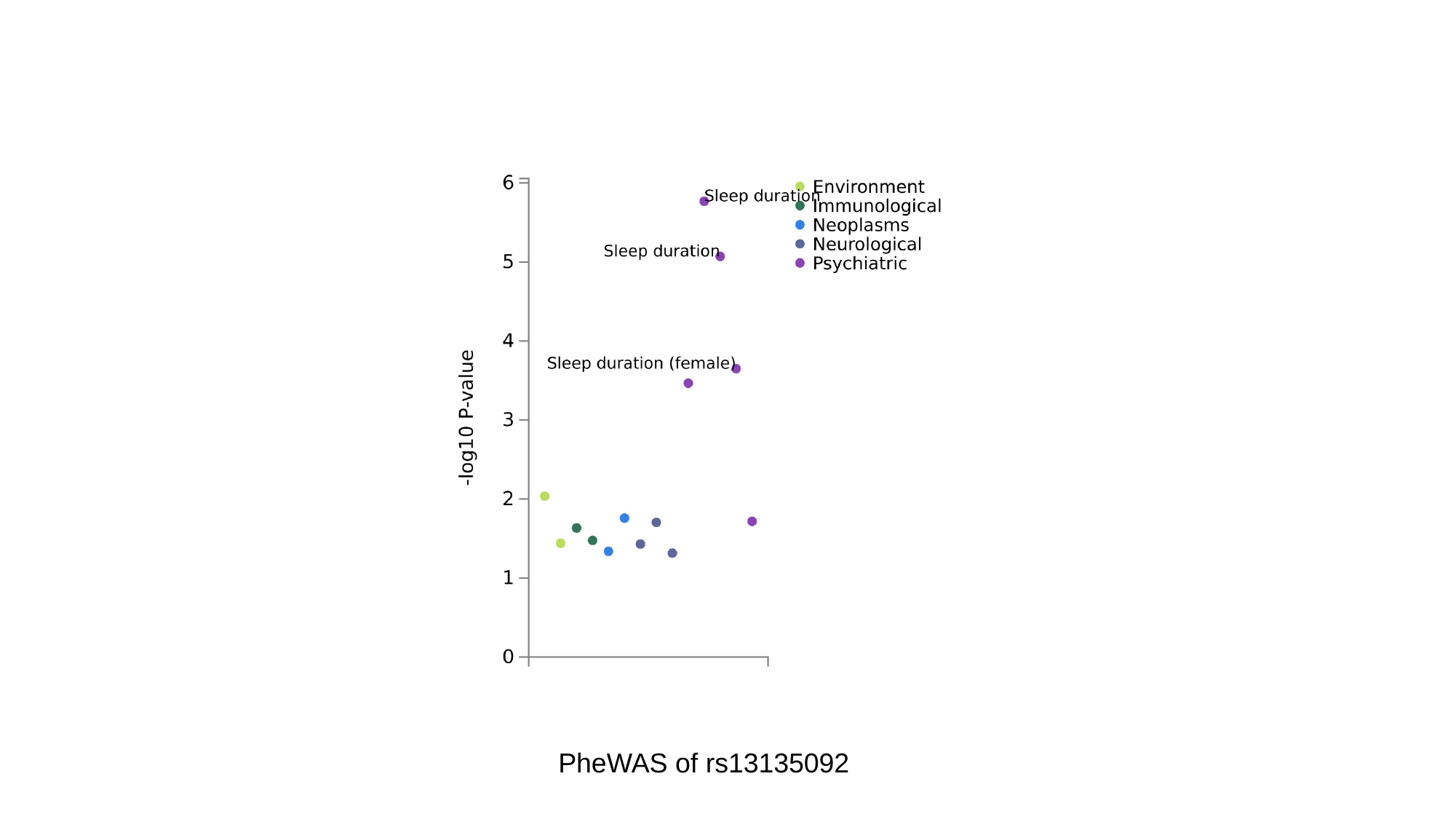

PheWAS of rs13135092

### Slide 4
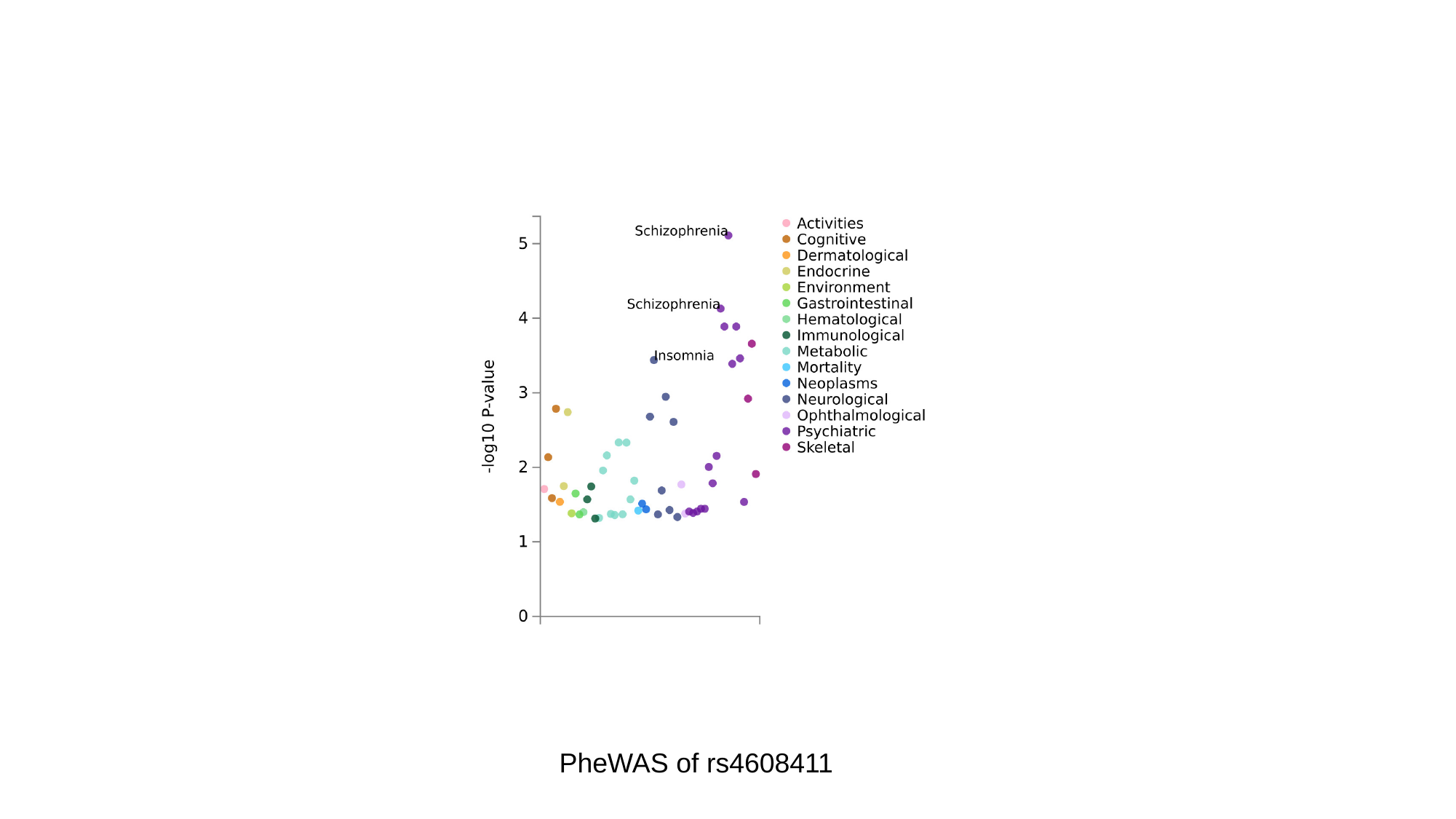

PheWAS of rs4608411

### Slide 5
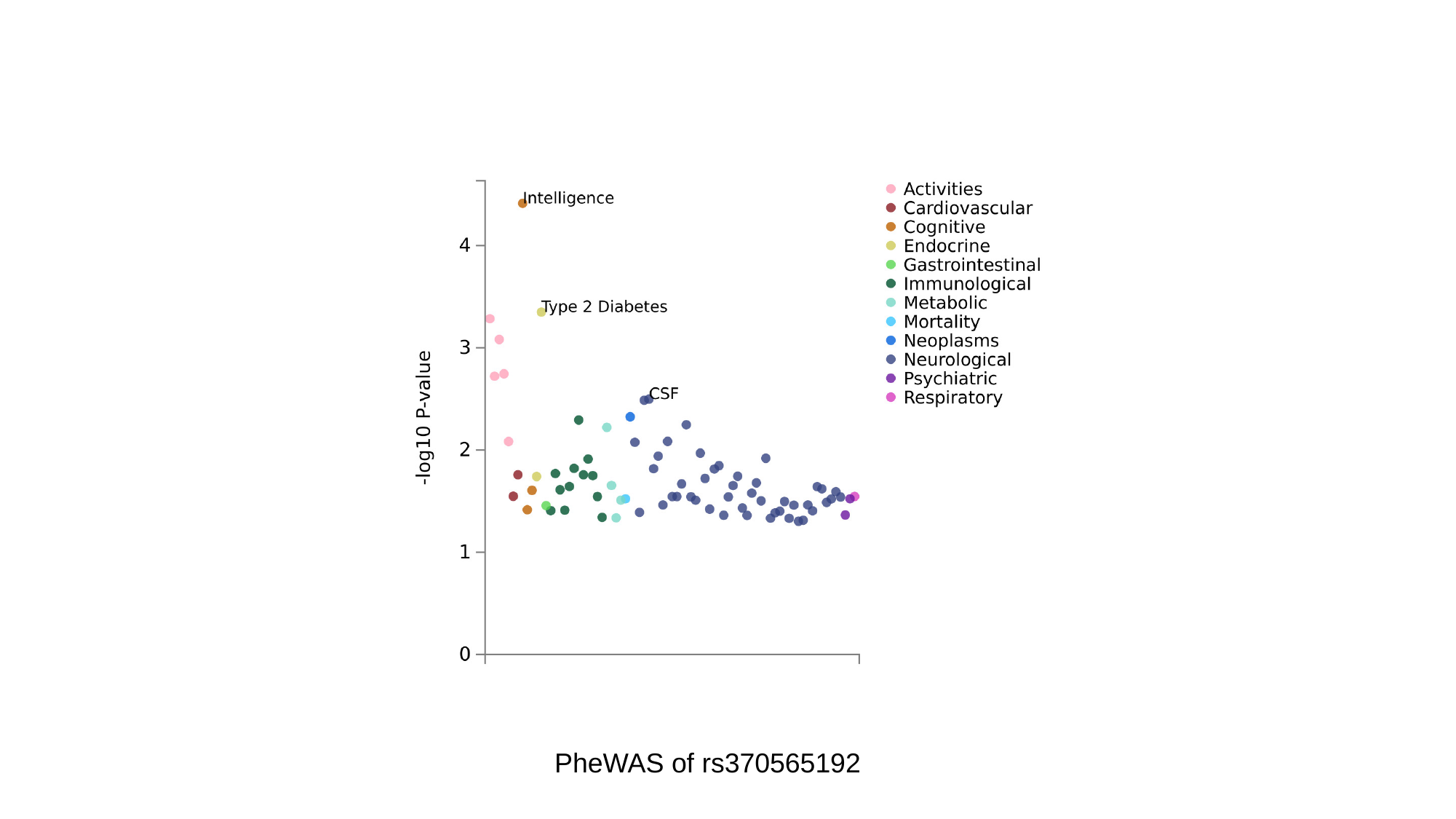

PheWAS of rs370565192

### Slide 6
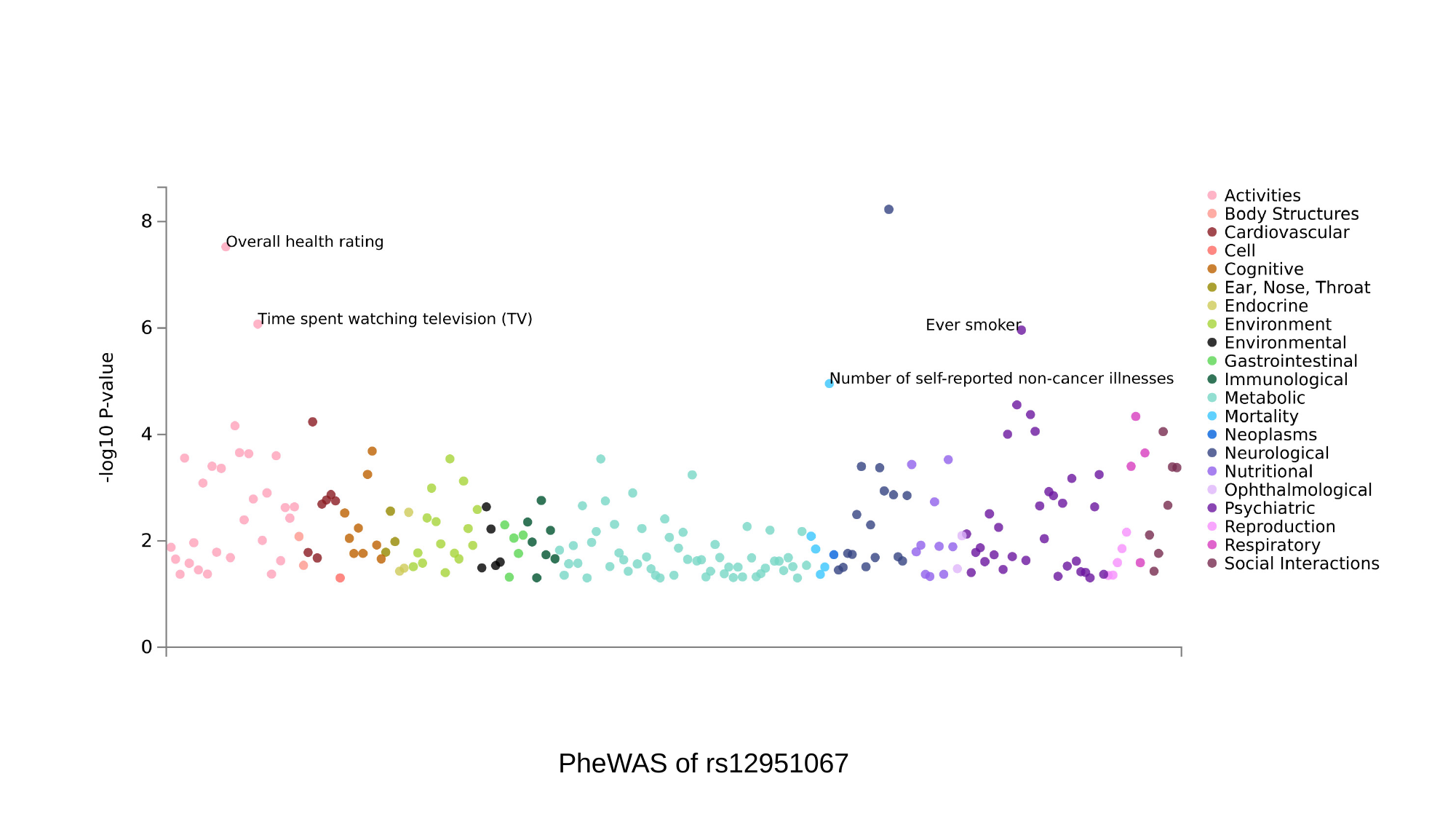

PheWAS of rs12951067
