## Supplementary Figures 4 for "A Genome-wide Association Study Identifies Novel Genetic Variants Associated with Neck or Shoulder Pain in the UK Biobank (N = 441,757)"

### Slide 1
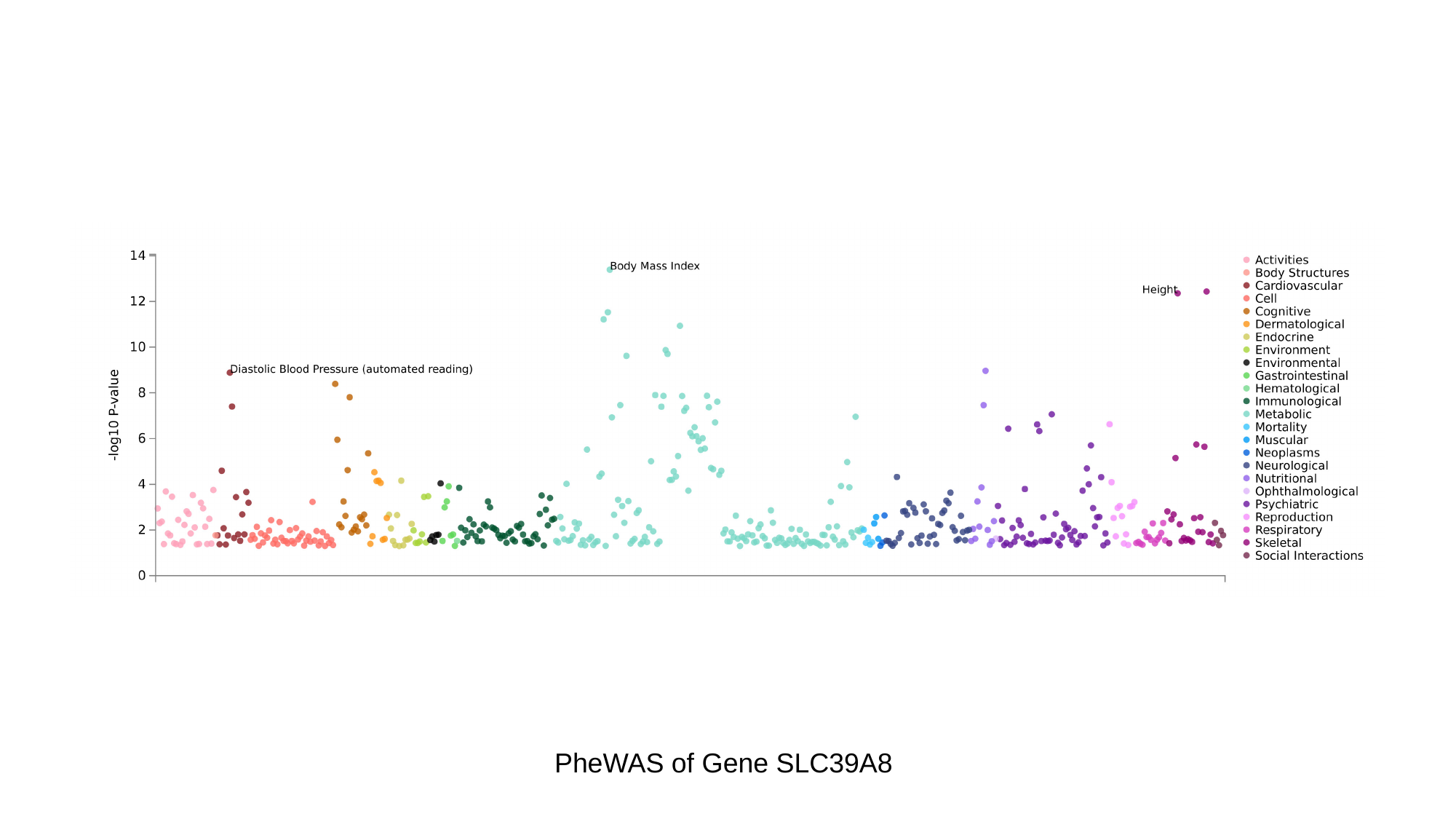

PheWAS of Gene SLC39A8

### Slide 2
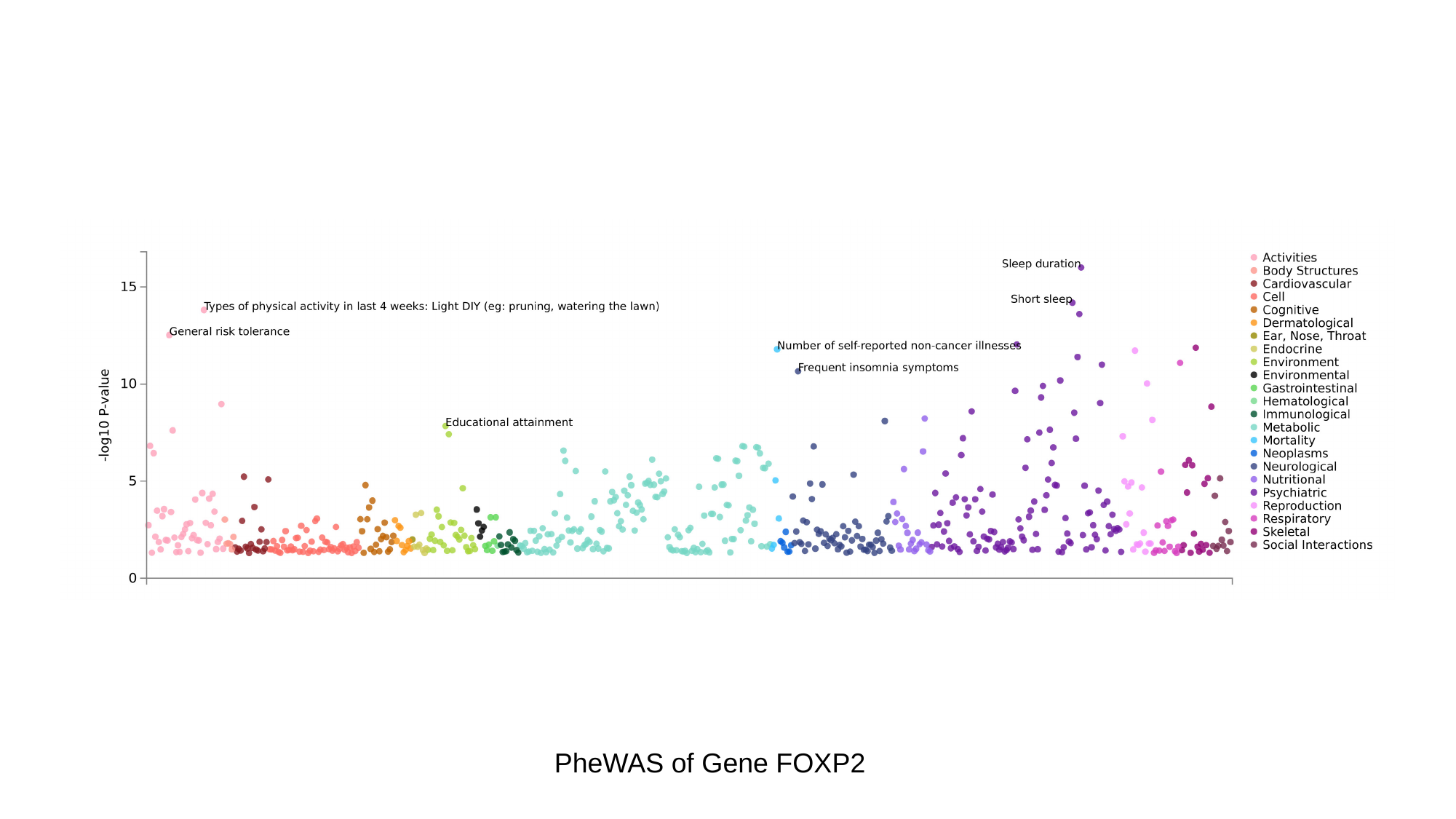

PheWAS of Gene FOXP2

### Slide 3
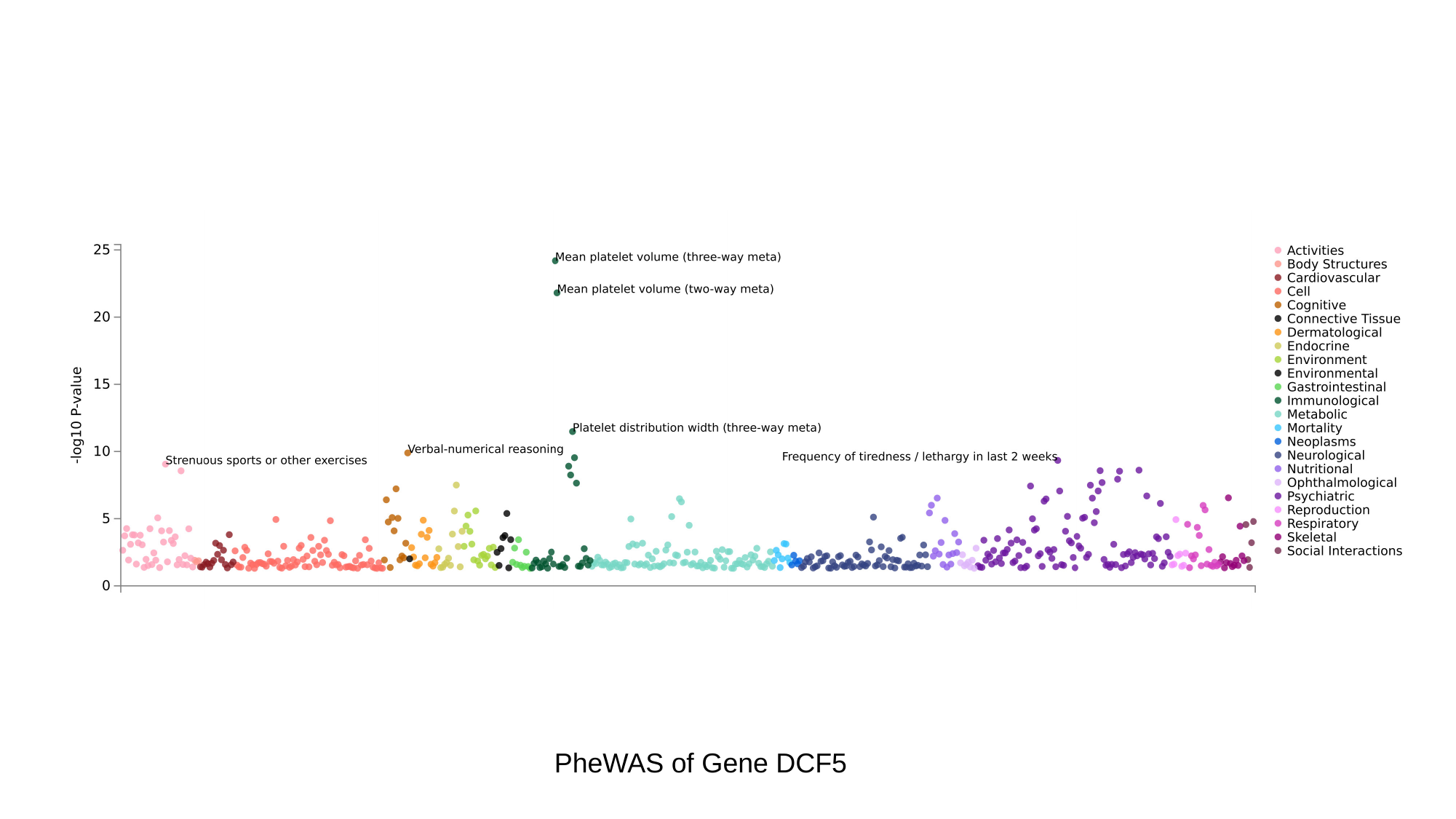

PheWAS of Gene DCF5

### Slide 4
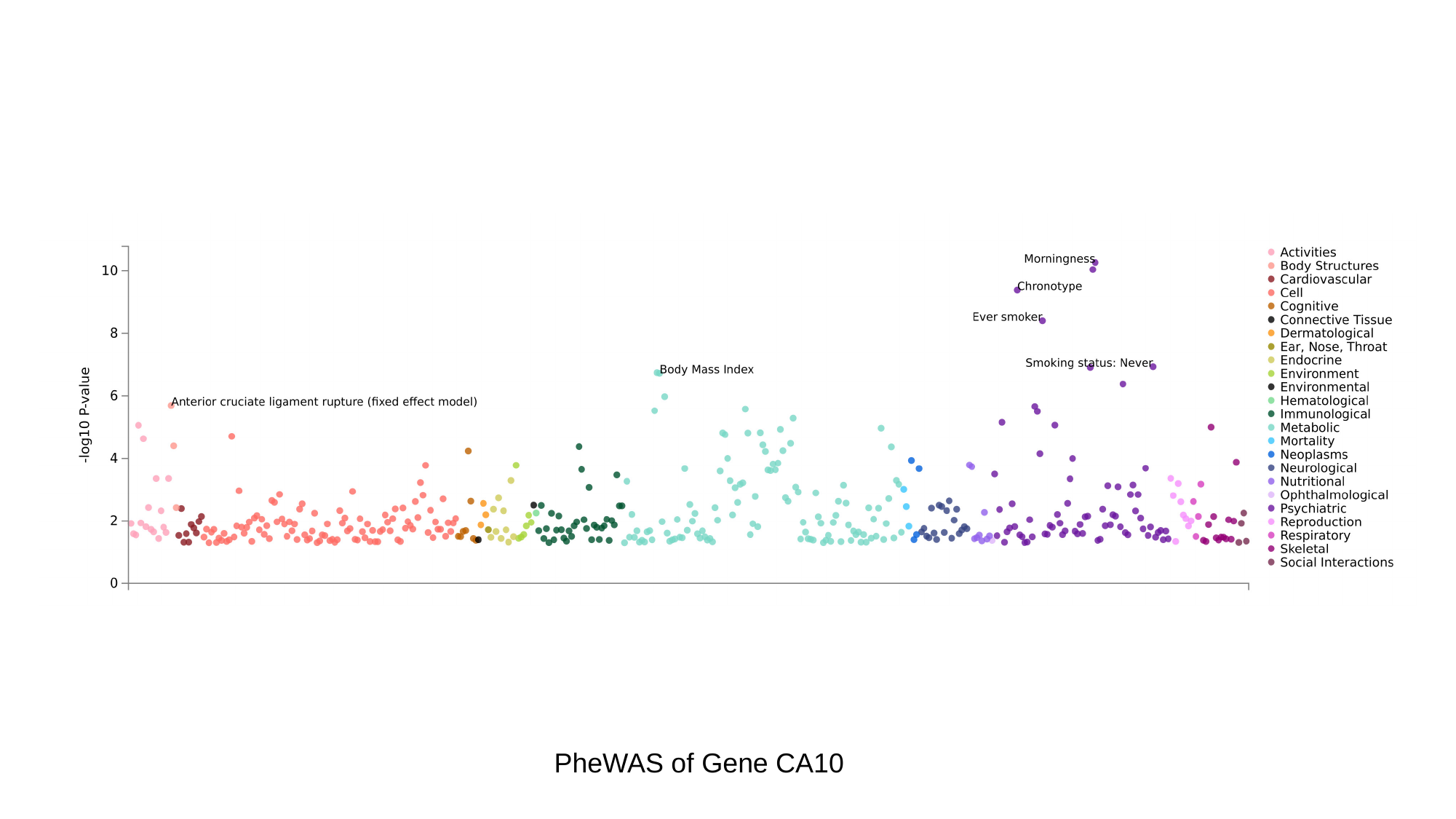

PheWAS of Gene CA10

### Slide 5
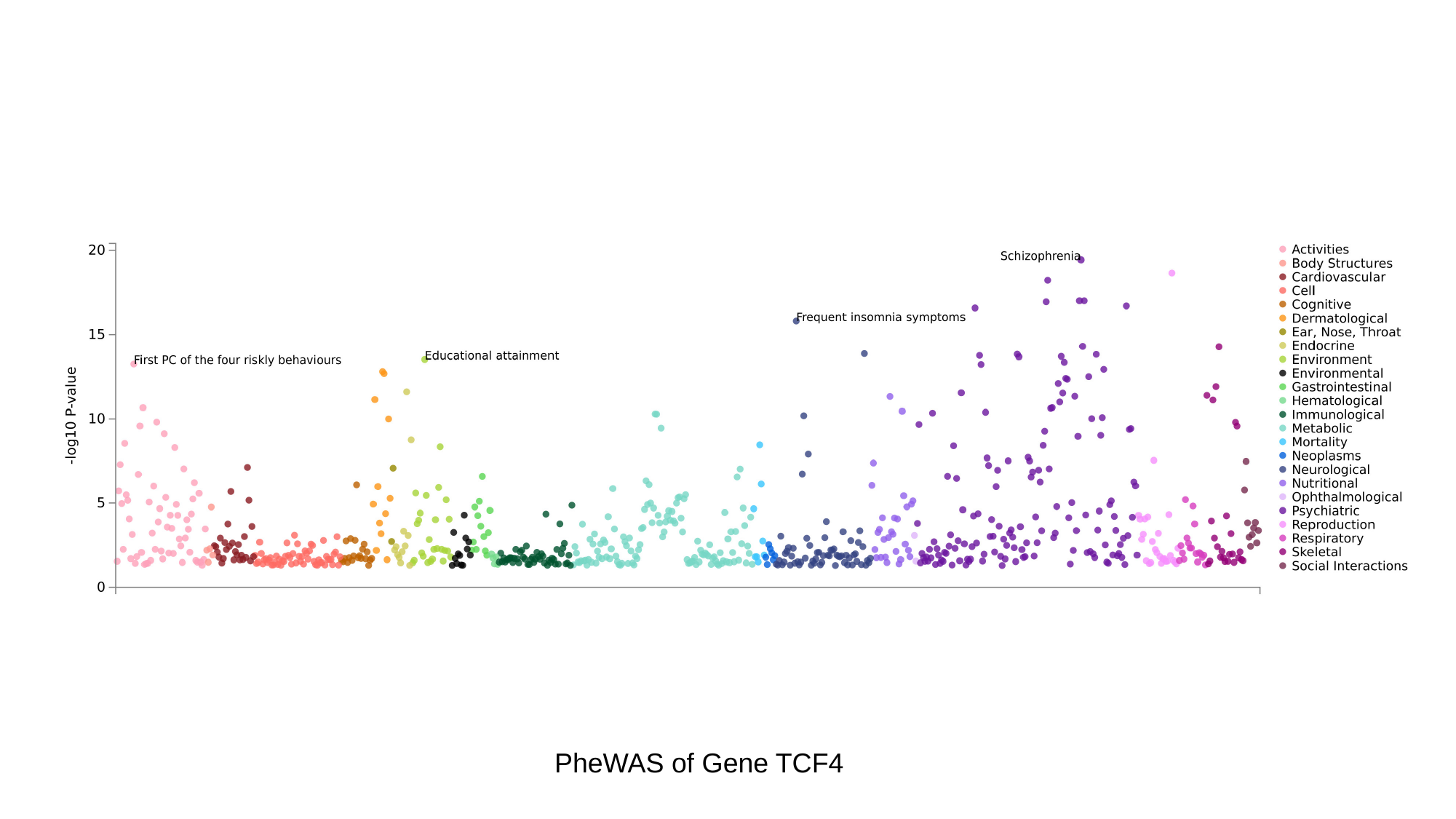

PheWAS of Gene TCF4
